# Genetic and environmental correlations between complex phenotypes differ by race/ethnicity and sex

**DOI:** 10.1101/2021.09.05.21263126

**Authors:** Michael Elgart, Matthew O. Goodman, Carmen Isasi, Han Chen, Paul S. de Vries, Huichun Xu, Ani W Manichaikul, Xiuqing Guo, Nora Franceschini, Bruce M. Psaty, Stephen S. Rich, Jerome I. Rotter, Donald M. Lloyd-Jones, Myriam Fornage, Adolfo Correa, Nancy L. Heard-Costa, Ramachandran S. Vasan, Ryan Hernandez, Robert C. Kaplan, Susan Redline, the Trans-Omics for Precision Medicine (TOPMed) Consortium, Tamar Sofer

**Affiliations:** Division of Sleep and Circadian Disorders, Brigham and Women’s Hospital, Boston, MA, USA; Department of Medicine, Harvard Medical School, Boston, MA, USA; Department of Epidemiology and Population Health, Albert Einstein College of Medicine, Bronx, NY, USA; Human Genetics Center, Department of Epidemiology, Human Genetics, and Environmental Sciences, School of Public Health, The University of Texas Health Science Center at Houston, Houston, TX, USA; Center for Precision Health, School of Biomedical Informatics, The University of Texas Health Science Center at Houston, Houston, TX, USA; Department of Medicine, University of Maryland School of Medicine, Baltimore, MD, USA; Center for Public Health Genomics, University of Virginia, VA, USA; The Institute for Translational Genomics and Population Sciences, Department of Pediatrics, The Lundquist Institute for Biomedical Innovation at Harbor-UCLA Medical Center, Torrance, CA, USA; Department of Epidemiology, University of North Carolina, Chapel Hill, NC, USA; Cardiovascular Health Research Unit, Departments of Medicine, Epidemiology, and Health Services, University of Washington, Seattle, WA, USA; Center for Public Health Genomics, University of Virginia School of Medicine, Charlottesville, VA, USA; Department of Preventive Medicine, Northwestern University, Chicago, IL, USA; Brown Foundation Institute of Molecular Medicine, McGovern Medical School, University of Texas Health Science Center at Houston, Houston, TX, USA; Department of Population Health Science, University of Mississippi Medical Center, Jackson, MS, USA; Boston University and National Heart Lung and Blood Institute’s Framingham Heart Study, Framingham, MA, USA; Department of Neurology, Boston University School of Medicine, Boston, MA, USA; Preventive Medicine & Epidemiology, and Cardiovascular Medicine, Medicine, Boston University School of Medicine, and Epidemiology, Boston University School of Public health, Boston, MA, USA; Department of Bioengineering and Therapeutic Sciences, University of California, San Francisco, CA, USA; Fred Hutchinson Cancer Research Center, Division of Public Health Sciences, Seattle, WA, USA; Department of Biostatistics, Harvard T.H. Chan School of Public Health, Boston, MA, USA

## Abstract

We developed novel closed-form estimators of genetic and environmental correlation coefficients. We applied them to estimate over 4,000 genetic and environmental correlations between multiple phenotypes in a diverse sample from the Trans-Omics in Precision Medicine (TOPMed) program. We found substantial differences in heritabilities, genetic, and environmental correlations of multiple phenotypes and phenotype-pairs between Black, Hispanic/Latino and White populations as well as between sexes. Finally, we quantified genetic and environmental correlations between phenotypic domains, each characterized by multiple phenotypes. Altogether we provide a novel, in-depth framework for examining relations among complex human phenotypes and their determinants.

## 2. Introduction

Both genetics and environment determine human phenotypes and the correlations between them [1, 2]. Correlations can arise due to multiple forms of causal relationships including common genetic and environmental determinants, (bi)directional causal associations and others. The correlations between phenotypes can reveal genetic architecture, help uncover gene functions and disease mechanisms, improve diagnosis and aid in therapeutic interventions [3]. Given appropriate data, phenotypic correlations can be decomposed into genetic and environmental components by estimating corresponding measures, such as genetic correlation [4].

Several studies have leveraged the data generated by large studies with genotyped individuals and multiple measured phenotypes (e.g. BioBank Japan (BBJ)[5] and UK Biobank (UKB)[6]), to estimate genetic correlations between various phenotypes [7–10]. Dozens of pairwise correlations between phenotypes were estimated and reported with the vast majority of participants being of the same race/ethnicity of either European or East Asian descent[9, 10] and of mixed genders. Race/ethnicity and gender, are complex constructs, associated with differences in phenotypic distributions between different subpopulations due, to some extent, to both underlying genetics and different environmental exposures [11–13]. The genetic background of a subpopulation manifests in allele frequencies, effect sizes, and more generally, genetic architectures, driven by both genetic ancestry composition of the subpopulation, and by behavioral, environmental and psychosocial exposures such as smoking, alcohol, nutrition, physical activity, and stress [14, 15], modifying the effect of genetic variants. While a handful of studies reported differences in genetic correlation across race/ethnic groups [16–18], these are limited in the number of studied phenotypes and race/ethnicities. Similar to race/ethnicity, while sex-specific heritabilities of complex phenotypes have been previously reported [19–21], sex-specific correlations between phenotypes have not been comprehensively studied.

The two main computational approaches that are used to estimate the genetic correlations are the genetic restricted maximum likelihood analysis (GREML)[22–24], which requires individual-level genotypes and is computationally challenging when analyzing datasets with thousands of individuals; and the linkage disequilibrium score regression (LDSC) [25], which uses genome-wide association study (GWAS) summary statistics. LDSC requires reliable GWASs, and can be inaccurate when there is genetic heterogeneity between the target sample and reference LD panel [26, 27] and thus cannot be used reliably for admixed or multi-ancestry analyses. Thus, both GREML and LDSC approaches are limited for datasets that include tens of thousands of genetically diverse individuals.

Here, we estimated the heritabilities, as well as genetic and environmental correlations between phenotypes in multi-ethnic as well as race/ethnicity- and sex-stratified analysis. First, we derived a closed form solution for the estimation of genetic and environmental correlation coefficients within the Haseman-Elston regression framework. Second, we applied the algorithm to study heritabilities and genetic correlations for 28 phenotypes in the Trans-Omics in Precision Medicine (TOPMed) program [28, 29] dataset, with large representation of White, Black and Hispanic/Latino populations. We then focused on the Hispanic/Latino population from the Hispanic Community Health Study / Study of Latinos (HCHS/SOL) cohort [30], and utilized available data on shared household (representing shared environmental exposure) and compared sex-specific genetic and environmental correlations across a larger panel of 61 phenotypes. Finally, we performed domain-level enrichment analysis to identify genetic and environmental correlations between phenotypic domains.

## 3. Results

### A Compendium of genetic correlations and heritabilities in the multi-ethnic TOPMed dataset

We developed a novel closed-form estimator for genetic correlations between phenotypes within the Haseman–Elston regression framework. We studied the algorithm in simulations and compared the estimates to those estimated using GREML algorithm [4] implemented in the GCTA software [31] (Supplementary Fig 1), matching the latter for accuracy and improving the speed ∼100-fold. We calculated phenotypic and genetic correlations between 28 phenotypes using 33,959 TOPMed individuals. Results are provided in Figure 1 and Supplementary Data file 1. There were 378 phenotype pairs in the dataset. Of these, 227 (∼60%) had significant phenotypic correlations *R*, defined as p-value < 0.05 and |*R*|>0.05 (Fig1A). Of these, 82 (36%) also had genetic correlations *ρ*_*k*_ with p-value<0.05 (Fig 1B, Fig 1D-inset). In contrast, out of the 151 phenotype pairs having |*R*| < 0.05 and p-value<0.05, 21 pairs (∼9%) had *ρ*_*k*_ with p-value<0.05 (for FDR corrected p-values see Supplementary Data file 1).

**Figure 1.**
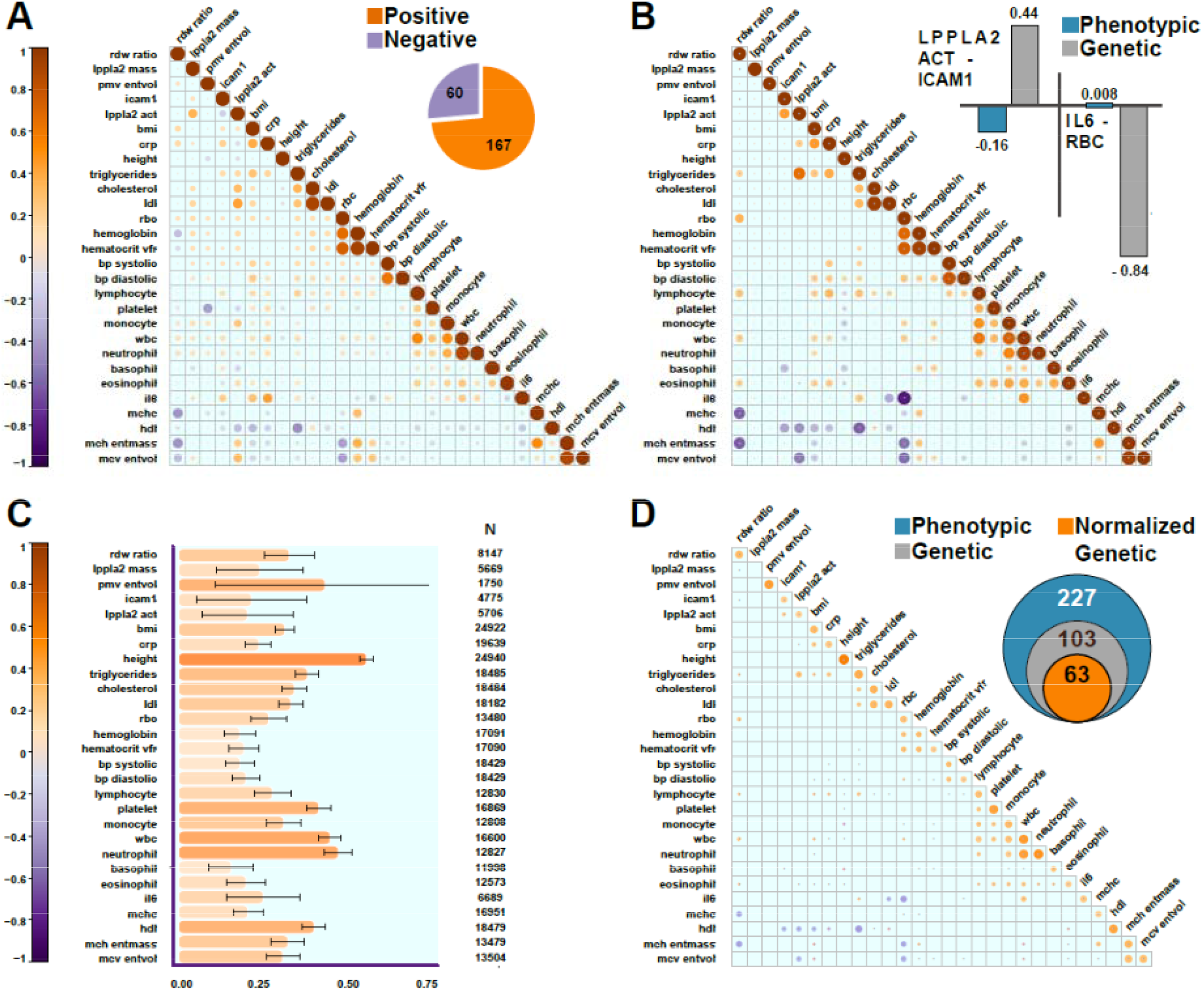
A third of observed phenotypic correlations between phenotypes in the combined TOPMed dataset have substantial genetic component. Correlation matrices where each column and row represent one of the 28 phenotypes in the TOPMed dataset and the intersection is the estimated correlation magnitude. Size and color of circle indicates the correlation strength; dark orange – positive, and dark purple – negative correlation. **(A)** Phenotypic correlations between the phenotypes. **Inset** – number of positive and negative correlated phenotype-pairs with p-value<0.05 and absolute phenotypic correlation above 0.05 **(B)** Estimated genetic correlations (*ρ*_*k*_) (shown only for phenotype pairs with p-value<0.05 between the phenotypes and |*ρ*_*k*_| > 0.05). **Inset** – examples of phenotypic and genetic correlation for two selected phenotypes where the absolute value of the genetic correlation is larger than that of the phenotypic correlation which complicates interpretability. **(C)** Estimated heritabilities for the studied phenotypes. **(D)** Normalized Genetic Correlations(*ρ*_*Nk*_) between the phenotypes (shown only for phenotype pairs with p-value < 0.05; and (*ρ*_*Nk*_) > 0.05). **Inset** – number of phenotype-pairs with phenotype, *ρ*_*k*_, and *ρ*_*Nk*_ with p-values < 0.05 and absolute value of at least 0.05 in this dataset.

Our results agree well with previous reports. For example, we estimated a genetic correlation of *ρ*_*k*_ = 0.24 between body-mass index (BMI) and triglycerides while Bulik-Sullivan et al. estimated it at 0.26[10] and Cadbi et al. at 0.2 [27]. Similarly, we estimated *ρ*_*k*_ (genetic correlation) between C-reactive protein and high-density lipoprotein (HDL) to be -0.24, while Ligthart et al. reported it at -0.29 [32]. We also identified new and potentially clinically-relevant genetic correlations. For example, we estimated a significant *ρ*_*k*_ = 0.44 between lipoprotein-associated phospholipase A2 and Intercellular adhesion molecule-1 which may important for the study of coronary artery disease biology; and multiple novel genetic correlations between white blood cell types and blood pressure, that are consistent with literature implicating a biological association between inflammation and hypertension [33, 34]; Supplementary Table 2).

We next estimated heritabilities for all the phenotypes in our multi-ethnic dataset (Fig 1C; Supplementary Table 3). The most highly heritable phenotypes are height (Fig 1C; ∼0.56) as well as multiple blood cell measurements such as neutrophil counts (Fig 1C; ∼0.47) and total white blood cell counts (WBC; Fig 1C; 0.45), similar to previous estimates [35, 36].

### Normalized Genetic Correlation integrates genetic correlations with heritabilities to uncover the direct contribution of genetics to the observed phenotypic correlations

Our results identified multiple instances where the genetic correlation coefficient is larger than the phenotypic correlation (see Fig 1b inset for examples), emphasizing that the genetic correlation coefficient is not directly related to the phenotypic correlation [37, 38], and may be appreciable, even in pairs with overall very low phenotypic correlation. We thus introduce the concept of “normalized genetic correlation” (*ρ*_*Nk*_), defined as the component of the observed phenotypic correlation *R* explained by genetics in the decomposition 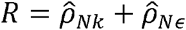,, where 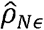, is the estimated normalizedresidual correlation.

We estimated *ρ*_*Nk*_ for all the pairs of phenotypes in our dataset (Fig 1D; Supplementary Table 4). Of phenotype pairs with genetic correlation with p-value<0.05, 61% had substantial 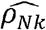, defined as 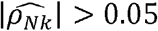, (Fig 1D-inset), corresponding to 23% of the phenotypic correlations with R >0.05. For most of the phenotype-pairs, 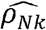 is much lower than 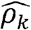 and is lower than the estimated *R* (as expected). For example, for BMI and C-Reactive Proein (CRP), 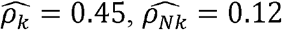, and 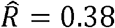. For HDL and triglycerides, 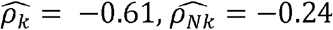, and 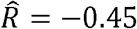.

### Genetic correlations between complex phenotypes and their heritabilities within race/ethnic groups

Out dataset includes 8,054 Black participants, 17,143 White participants and 8,762 Hispanics/Latinos (Supplementary Table 2). We estimated heritability, phenotypic, and genetic correlations within these race/ethnic groups and compared them to the multi-ethnic estimates (Figure 2; Supplementary Figure 2; Supplementary Data Files 1-4).

**Figure 2.**
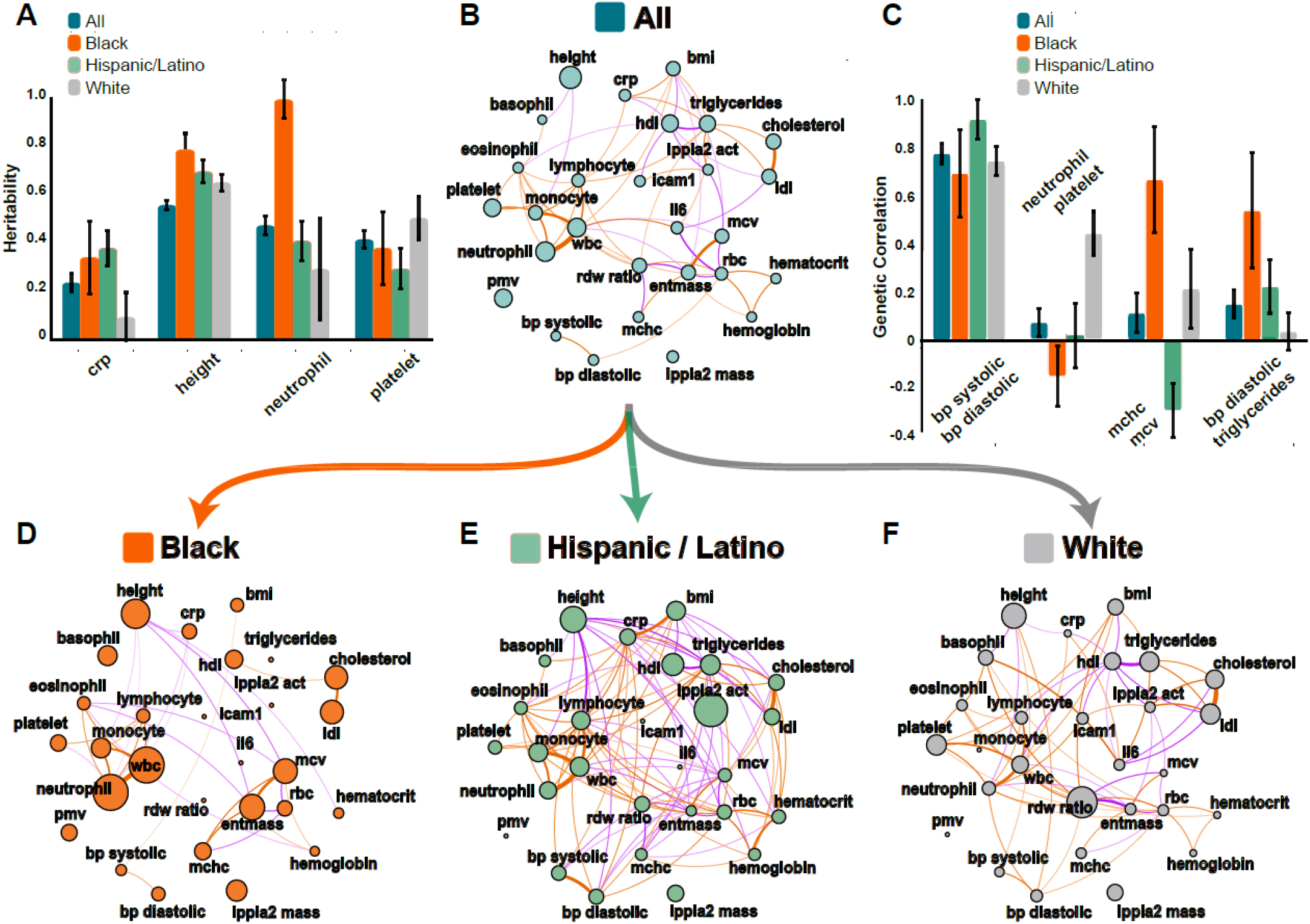
Some genetic correlations and heritabilities are race/ethnicity-specific. **(B, D-F)** Correlation plots where each phenotype is represented by a node and the correlations are represented by connections (edges) between nodes. The size of the node is proportional to the phenotype heritability. The thickness of the edge is proportional to the magnitude of correlation and the color represents direction: orange represents positive and purple negative correlation. **(A)** Examples of race/ethnic-specific heritabilities **(C)** Examples of race/ethnic-specific genetic correlations (*ρ*_*k*_) **(B)** Normalized genetic correlations (*ρ*_*Nk*_) between the 28 phenotypes in the combined TOPMed dataset (p-value < 0.05; |*ρ*_*Nk*_| > 0.05) **(D, E, F)** Normalized genetic correlations (*ρ*_*Nk*_) between the 28 phenotypes in the race/ethnic-specific subsets of the TOPMed dataset - Black (**D**, orange), Hispanic/Latino (**E**, marine) and White (**F**, grey).

While some phenotypes, such as HDL and eosinophil counts, have similar heritabilities across race/ethnicities (Fig 2A, D-F), many other heritabilities vary by race/ethnicity. For example, CRP is similarly heritable in the Blacks and Hispanics/Latinos (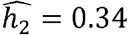 and 0.38 respectively), but much less so in Whites 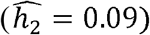. In contrast, neutrophil counts are very heritable in Blacks 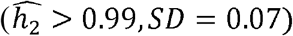, but are less so in Whites 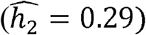 and Hispanics/Latinos 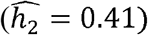.

Similarly, multiple phenotype-pairs such as neutrophil and total WBC counts, and systolic and diastolic blood are similarly genetically correlated across race/ethnic groups (Fig 2C, D-F). However,many other *ρ*_*k*_ differ by race/ethnicity (Fig2 C, D-F). For example, neutrophil and platelet counts have 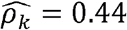 (p-value < 0.01) in Whites, but only 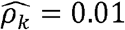 in Hispanics/Latinos, and 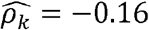 (p-value < 0.01) in Blacks (Fig2 C, D-F). Similarly, diastolic blood pressure and triglycerides (Figure 2C) are strongly genetically correlated with 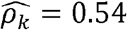 in Blacks but have 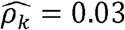 in Whites.

Overall, we detect 63 phenotype-pairs with |*ρ*_*Nk*_| > 0.05 and p-value<0.05 in the combined multi-ethnic TOPMed dataset while detecting only 33 such pairs in the Black population, 105 in the Hispanic/Latino population and 69 in the White population. Supplementary Figure 2A visualizes the overlap between these correlations across race/ethnic groups (FDR corrected p-values are provided in Supplementary Data files).

### Both genetics and shared-household drive phenotypic correlations in Hispanics/Latinos

We studied genetic correlations among a larger panel of 61 phenotypes in n=7,678 Hispanics/Latinos from the HCHS/SOL [30, 39]. The phenotypes represent 11 phenotypic domains: diabetes, cardiovascular disease, blood pressure, kidney function, lipids, lung function, sleep, anthropometrics, iron, RBC (red blood cells) and WBC (Fig 3, Supplementary Table 1). HCHS/SOL also has information about household sharing between participants, allowing for estimation of both genetic and environmental correlations between phenotypes.

**Figure 3.**
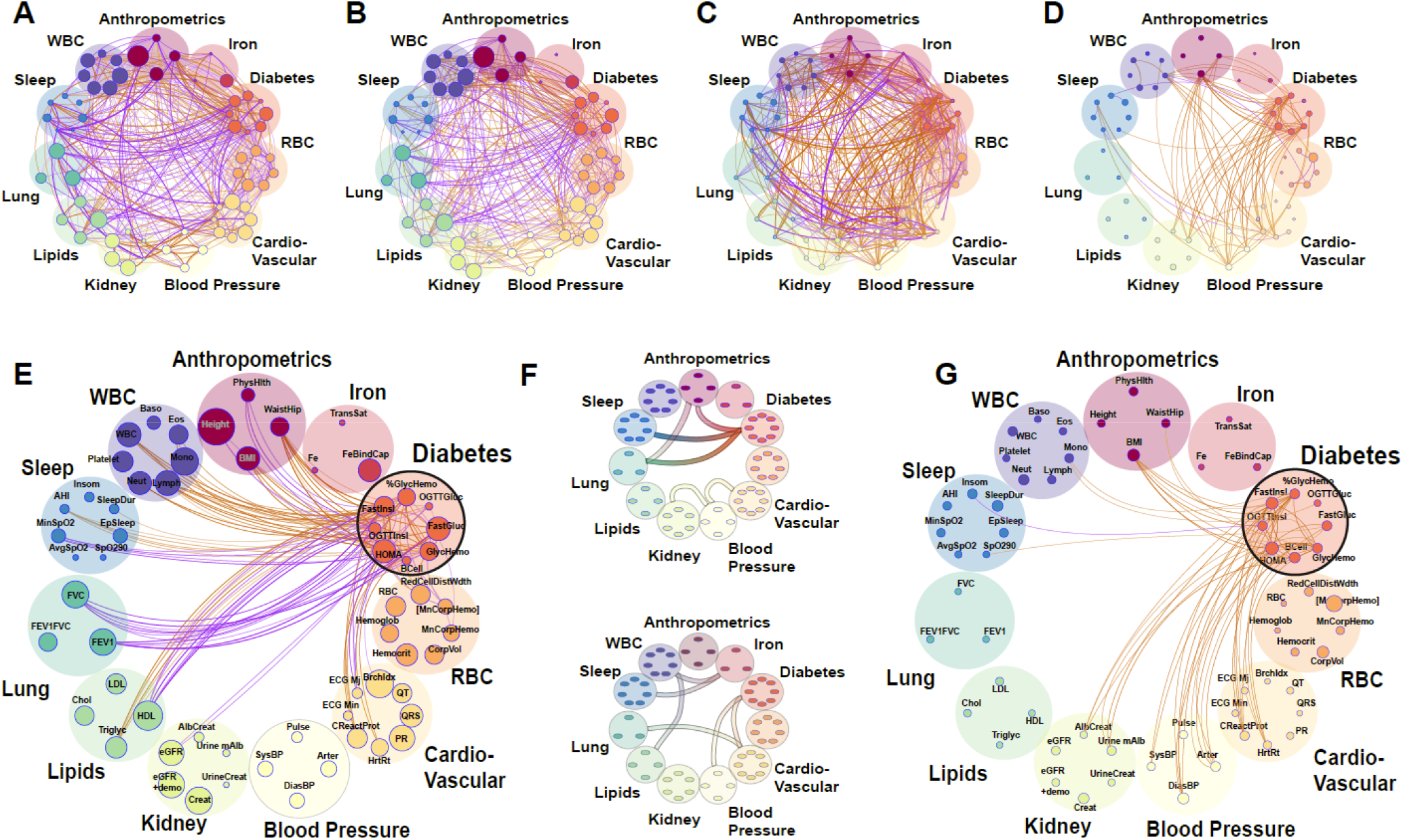
Genetics and shared Household factors contribute to associations between phenotypes in Hispanics/Latinos. **(A-F)** Correlation plots between the 61 phenotypes in the TOPMed HCHS/SOL dataset. Each phenotype is represented by a node (colored small circles) with the size of the circle proportional to the phenotype heritability. The correlations are represented by connections (edges) between nodes (phenotypes). The nodes are grouped into phenotypic domains (colored semi-transparent circles labelled Anthropometrics, Iron, etc.). The thickness of the edge is proportional to the strength of correlation and the color represents magnitude: orange represents positive and purple negative correlation. **(A)** Genetic correlations (*ρ*_*k*_) between the 61 phenotypes (p-value < 0.05; |*ρ*_*k*_| > 0.05) **(B)** Normalized genetic correlations (*ρ*_*Nk*_) between the 61 phenotypes (p-value < 0.05; |*ρ*_*Nk*_| > 0.05) **(C)** Household correlations (analog of genetic correlation but for household data; see Materials and Methods) between the 61 phenotypes (p-value < 0.05; |***ρ***_***H***_| > 0.05) **(D)** Normalized household correlations (*ρ*_*Nh*_) between the 61 phenotypes (p-value < 0.05; |*ρ*_*Nh*_| > 0.05). **(E+G)** A focused look on the normalized genetic correlations for the contribution to the diabetes phenotype-domain of genetics (**E**) and shared household (**G**). Only the correlations for the diabetes phenotype-group are displayed. **(F)** Connections represent significantly enriched correlations between the phenotypic domains. (F-Top) represents genetic correlations and (F-bottom) Household correlations.

We estimated *ρ*_*k*_ and *ρ*_*Nk*_ in conjunction with the equivalent household correlation measures *ρ*_*h*_ and *ρ*_*Nh*_ for all the 1,830 pairs of phenotypes (figure 3). Out of the 1,830 phenotype-pairs, 1007 (or ∼55%) have 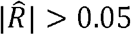 (Supplementary Data file 5). Of these, 403 (∼40%) also have 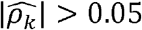 (Fig 3A) and 412 (∼41%) have 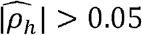 with p-value<0.05. Of the genetically correlated pairs, 343 phenotype-pairs (∼34% of all phenotypic correlations) have |*ρ*_*Nk*_| greater than 0.05. Of the 412 phenotype-pairs with significant 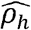, only 91 are of |*ρ*_*Nh*_| > 0.05 (∼9% of all phenotypic correlations). Unlike heritabilities, the estimated variance components for household are quite low for all phenotypes (Supplementary Data File 5), resulting in low *ρ*_*Nh*_ compared to *ρ*_*Nk*_. An interesting contrast between the genetic and household correlation is in the diabetes domain (Figure 3E, G). The diabetes domain has strong household correlations with blood pressure domain phenotypes (but no observed genetic correlations) and has strong genetic correlations with lung and lipid domain phenotypes (but no observed household correlations).

### Shared genetic and environmental basis between phenotypic domains via group enrichment analysis

The genetic correlations are distributed non-uniformly with regard to the phenotypic domains. Domain enrichment analysis showed a strong enrichment of the number of intra-group correlations for all the 11 group-phenotypes (Supplementary Data File 5). Figure 3F visualizes the significant domain-level correlations (p-value < 0.05) estimated in enrichment analysis. Significant inter-group genetic correlations are estimated between the blood pressure domain and the kidney and cardiovascular domains; between the anthropometrics domain and the lung and diabetes domains; and between the diabetes domain and the anthropometrics, lung and sleep domains (Fig 3F-Top). Domain-level correlations due to shared household do not mirror the genetic ones (Fig 3F-Bottom vs Top). Specifically, we see that shared household affects the correlations between the diabetes and the blood-pressure and cardiovascular domains; the iron domain and the sleep and WBC domains; and the cardiovascular and lung domains (Fig 3F-Bottom).

### Heritabilities as well as genetic and environmental correlations between complex human phenotypes differ across sexes

To obtain sufficient sample size, we expanded our analysis to include all genotyped participants in the HCHS/SOL cohort using imputed genetic data, for a total of 12,565 participants: 5,175 males and 7,390 females. Results are provided in Figure 4, with the number of significant correlations and overlaps between sexes visualized in Supplementary Figure 4. Supplementary Figure 5 and Supplementary Table 5 further provide additional results from sex-stratified genetic correlation analysis in TOPMed Whites. Overall, there were notable differences between both genetic and environmental correlations between the sexes. For example, the phenotype pair BMI and AHI (Apnea Hypopnea Index) had 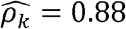 with p-value < 0.01 in men but 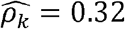 with p-value = 0.02 in women; and WaistHip (waist to hip ratio) and GlycHemo (Glycosylated Hemoglobin in SI units) had 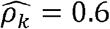 with p-value < 0.01 in females but 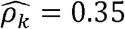 with p-value < 0.01 in males. Similarly, the household correlation for HOMA (HOMA-IR index of Insulin Resistance) and Height is 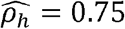 with p-value < 0.01 in men but 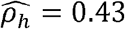 with p-value < 0.01 in women.

**Figure 4.**
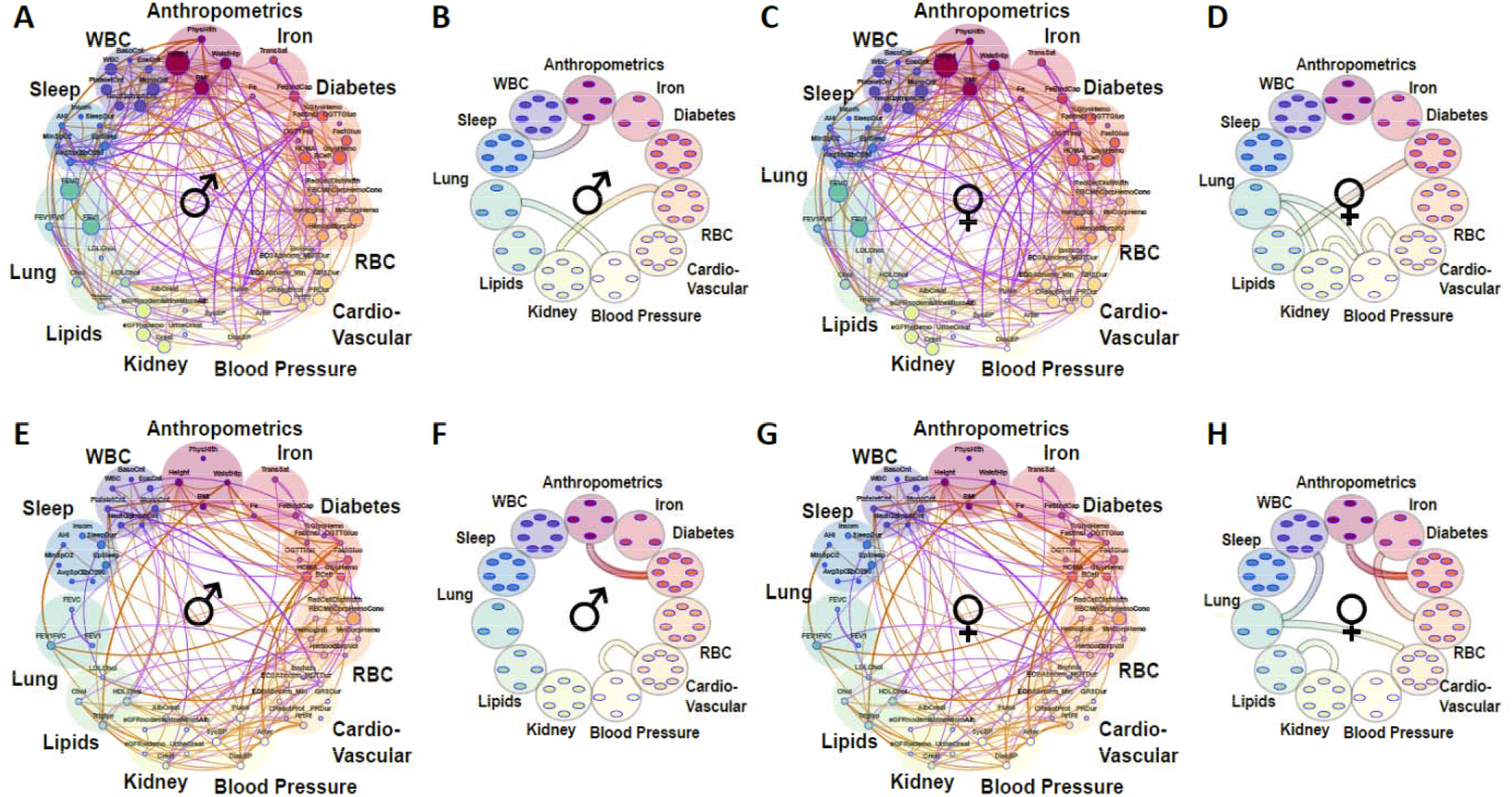
Sex differences in genetic and environmental correlations and heritabilities. **(A, C, E, G)** Correlation plots where each phenotype is represented by a node and the correlations are represented by connections (edges) between nodes. The size of the node is proportional to the phenotype heritability. The thickness of the edge is proportional to the strength of correlation and the color represents magnitude: orange represents positive and purple negative correlation. Shown are genetic correlations (*ρ*_*k*_) between the 61 phenotypes in the extended HCHS/SOL dataset (p-value < 0.05; |*ρ*_*k*_| 0.05). Correlations and heritabilities as measured in males **(A, E)** and females **(C, G). (B, D, F, H)** Significantly enriched correlations between the phenotypic domains. The top panels represent genetic correlations **(A-D)** and the bottom panels **(E-H)** represent the environmental correlations.

Multiple differences can be observed at the domain level (Figure 4 B+D, F+H). For example, while the correlations between Kidney function and RBC (red blood cells) are predominantly genetic in males (Figure 4 B vs D), and do not reach significance threshold in females, the correlations between Anthropometrics and Diabetes are predominantly due to shared household regardless of sex (Figure 4 F vs H). The same correlations can be differentially driven in sex-dependent manner as Blood Pressure interacts with Cardio-vascular domain through genetics in females but through environment in males (Figure 4 D vs F).

## Discussion

We developed and implemented a novel, computationally efficient framework to estimate genetic and environmental correlations between phenotypes, and their contributions to the overall phenotypic correlation. We systematically interrogated heritabilities and genetic correlations between all phenotype-pairs of 28 blood pressure, lipids, blood counts, inflammation, and anthropometrics/demographic phenotypes from individual-level TOPMed dataset totaling 33,959 individuals, further stratified by race/ethnic group. We then focused on Hispanic/Latino participants from the HCHS/SOL cohort and estimated heritabilities and genetic and environmental (due to shared household) correlations between 61 phenotypes from 11 domains (diabetes, cardiovascular, blood pressure, kidney, lipids, lung, sleep, anthropometrics, iron, RBC, and WBC), further stratified by sex. Finally, we identified enriched genetic and environmental correlations at the domain level.

The genetic correlations for many of the phenotype pairs in our dataset have never been reported in the literature, and previous reports typically evaluated correlations in ancestrally uniform cohorts mostly without Black and Hispanic/Latino participants. The estimated heritabilities agree well with previously reported values from large population-based studies, and are slightly lower than heritability estimates from twin studies [5, 22, 24, 35, 40]. A recent report showed that higher heritability can be recovered from WGS studies by separating genetic variants into bins of allele frequencies, and constructing multiple genetic relationship matrices within those bins [41]. It is a topic of future research to study genetic correlations in these settings. Notably, our framework allows for multiple correlation matrices, enabling such investigation.

In our primary TOPMed dataset (as well as in several previous reports [37, 38]), there are multiple instances where the estimated *ρ*_*k*_ is greater than the phenotypic correlation *R* (see Fig 1b - inset). For example, while we estimated a modest phenotypic correlation of 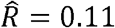 between CRP and lymphocyte counts, the corresponding genetic correlation estimate was 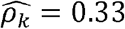. Thus, it is impossible to infer what fraction of the observed correlation is driven by genetics. We therefore introduce the concept of “normalized genetic correlation coefficient” (*ρ*_*Nk*_) as the component of the observed phenotypic correlation explained by genetics. This concept allows for identification of phenotype-pairs where genetics is a large contributor to the observed correlation. Of phenotype pairs with genetic correlation |*ρ*_*k*_| > 0.05, only ∼17% of all phenotype-pairs have “substantial” normalized genetic correlations, defined here as |*ρ*_*Nk*_| > 0.05 (Fig 1D-inset). For CRP and lymphocytes, 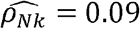, suggesting that the phenotypic correlation between these phenotypes is mainly driven by genetics (0.09 out of 0.11, which is 81%).

We were also able to estimate genetic and environmental correlations in understudied Hispanics/Latinos from the HCHS/SOL. The proportions of phenotype-pairs with significant phenotypic, genetic, and normalized genetic correlations, are overall similar to those in TOPMed, despite the expanded panel of phenotypes. We were also able to identify ∼22% of phenotype-pairs having |*ρ*_*h*_| > 0.05 with p-value < 0.05, but only ∼4% had normalized household correlation |*ρ*_*Nh*_| > 0.05, due to the overall low phenotypic variance explained by household environment. Shared household may contribute to the correlation between phenotypes by environmental causes of these phenotypes, e.g. nutrition, indoor pollutants, hygiene and other lifestyle habits, etc. We found that the association of diabetes phenotypes with lipids and lung functions is largely genetic, while the diabetes phenotypes’ associations with kidney function and blood pressure is mostly driven by shared environment. Future work is needed to study the implication of these results for treating complex cardiometabolic conditions.

In TOPMed, we studied heritabilities and genetic correlations stratified by self- or study-reported race/ethnicity. Some of the correlations and heritabilities differ across groups. This is not surprising given previous reports of differences in phenotypic distributions and disease prevalence across race/ethnicities [11–13]. For some phenotypes, the heritabilities were higher when estimated in each race/ethnic group separately. For example, while the estimated heritability for height in the combined dataset is 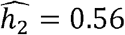, it was 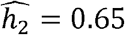 in Whites (similar to previous reports of ∼0.6-0.62 for individuals of European ancestry from TOPMed and UK Biobank [35]), 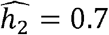 for Hispanics/Latinos and 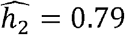 for Blacks. Different heritability estimates between combined and stratified models are likely due to more precise modelling of variances in the stratified models.

Similarly, when studying genetic correlations, we found fewer phenotype pairs with significant normalized genetic correlations in the combined TOPMed dataset (63 pairs) than in either of the comprising White and Hispanic/Latino race/ethnic groups (69 and 105 accordingly), despite lower sample sizes, but not in Blacks (33 pairs), and the correlations do not entirely overlap. The results of the combined dataset in Figure 1 likely represent the “consensus” genetic relationships of the phenotype-pairs, e.g. the pairs that behave consistently in the majority of the constituent race/ethnic groups. Interestingly, the Black group had the lowest number of significant genetically-correlated phenotype-pairs which may be due to the high genetic variability of African genetic ancestry (which is substantial in U.S. admixed Blacks) combined with low sample size and thus lower power [41].

In the HCHS/SOL cohort, where 61 phenotypes were available, we performed domain-level enrichment analysis akin to pathway enrichment analysis in gene expression studies. We defined domains as sets of phenotypes that capture similar underlying “latent” phenotypes, with the limitations that groups of phenotypes assigned for the same domain may still capture complex underlying biology, i.e. are not measures of exactly the same latent phenotype (e.g. insomnia and mean oxygen saturation during sleep, while can be correlated in individuals with obstructive sleep apnea, may also capture different pathophysiological disorders). While more study is needed, domain analysis should be less sensitive to individual variation in any particular phenotype. Interestingly, diabetes-domain correlations at the genetic level are enriched for sleep, anthropometry, and lipids domains, but at the environmental level, they are enriched for blood pressure and cardiovascular domains.

We performed sex-stratified analysis of genetic correlation in the Hispanics/Latinos, and found some sex differences. Overall, there were 32% phenotype-pairs with significant genetic correlations unique to females and 19% unique to males, and 23% and 15% with significant normalized genetic correlation. Similarly, there were 37% and 20% of pairs with significant household correlations in females and males accordingly but only 9% and 7% with significant normalized household correlation (Supplementary Figure 4 C-H) which was also generally weaker. Multiple correlations between phenotypic domains were also different between the sexes.

In interpreting household correlations, we note that much like race/ethnicity in its relationship to genetic ancestry, gender is a social construct that is related to sex, but is heavily influenced by social practices and norms. Environmental correlations may differ between genders due to sociocultural differences between them. Here, we were not able to assess socio-cultural contributions related to gender roles to the household correlations, and we refer to “sex” rather than “gender”, acknowledging that drivers of some of the estimated quantities are in fact gender, and not sex, based.

A specific strength of our study is the use of high-quality phenotypic and genotyping data from the diverse multi-ethnic TOPMed program. Further, all studies combined together are population-based cohort studies, reducing the likelihood of selection and other biases arising in studies following selected populations, such as case-control studies. Other strengths are the investigation of a large panel of phenotypes, evaluation of both genetic and environmental correlations, stratification by both race/ethnicity and sex, and the domain-level enrichment analysis. Nevertheless, this study also has a few limitations. For example, larger sample sizes, especially within stratified race/ethnic or sex groups, would enable stronger inferences. The use of self- or study-reported race/ethnicity rather than strata defined by genetic ancestry is also somewhat problematics. Still, we chose to proceed with these groupings, first, to reflect on currently used population groups in medical research, and second, because, given the use of Hispanics/Latinos who by definition, are admixed and cannot be represented by any detailed composition of genetic ancestry, there is no natural grouping that is based on genetic ancestry. As the field of genetic medicine grapples with the use of genetic ancestry and social definition of race/ethnicity [42], it would be important to re-consider models for genetic correlation analyses. Another limitation is imperfect definitions of phenotypic domains, that may not accurately capture underlying pathophysiology.

In summary, our work uncovered novel genetic and environmental correlations between phenotypes including differences by race/ethnicity and sex/gender. Future work includes the application of approaches from graph analysis field such as Gaussian Graphical Models [43, 44] [45] to discover directionality and causality; utilizing genetically correlated phenotypes to improve polygenic risk prediction models [46]; and studying genetic correlations by categories of genetic variants to capture the contributions of rare variants.

## 4. Methods

### 2.1. Statistical models

#### Statistical model when genetic relatedness in the only modeled source of correlation

Consider the linear model

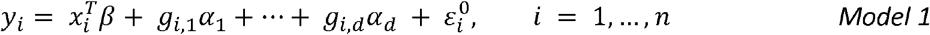

in which the quantitative outcome *y*_*i*_ is modelled by a regression on covariates *x*_*i*_ and the additive effects of *d* genetic variants *g*_1_,…,*g*_*d*_; and 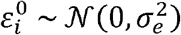 are normally distributed errors across *n* participants. Assuming that the genetic variants are independent random variables, each centered and scaled to have mean 0 and variance 1, the mean and variance of *y*_*i*_ are

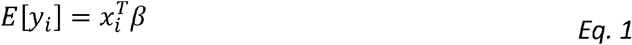

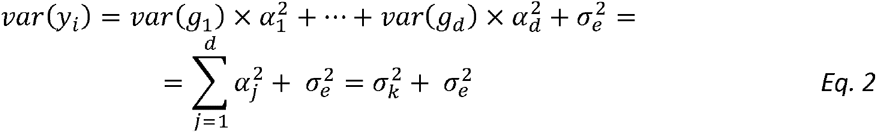

Here, 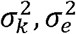 are the genetic and error variance components. Accordingly, narrow-sense heritability, which is the proportion of trait variance that is due to additive genetic factors is:

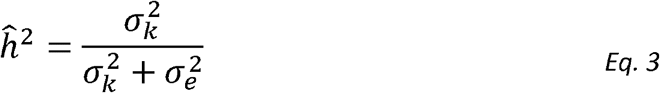

To model genetic correlation, we extend *Model 1* into a two-trait model. For person *i*:

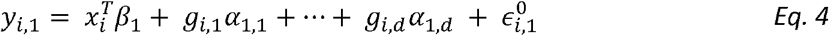

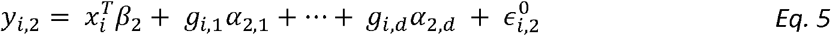

with error terms 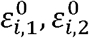, satisfying 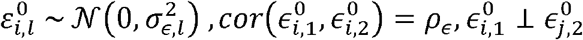 for *k* ∈ {1,2},*i* = 1,..,*n*.

Thus, the errors of the same person may be correlated for the two traits, but for different traits the error of person *i* is independent of the error of person *j*. Consider the covariance between the two traits, again while making the simplifying assumption of independence between genetic variants:

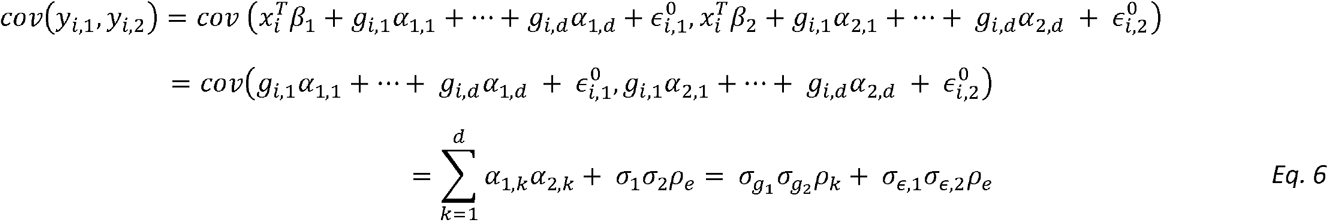

where *ρ*_*k*_ is the correlation between the genetic effect of the *k-th* variant on the two outcomes, and *ρ*_*e*_ is the correlation between the residual errors of the two outcomes. Note that the transition to using *ρ*_*k*_ at the final step treats the vectors of causal genetic effects *α*_1_ = (*α*_1,1…_*α*_1,*d*_)^*T*^, *α*_2_ = (*α*_2,1…_*α*_2,*d*_)^*T*^ as random variables with mean 0, i.e., 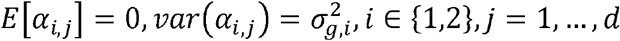.

Noting that for individuals *i,l, cov*(*g*_*i,j*_, *g*_*l,j*_) = *k*_*i,l*_ the probability of the two individuals sharing the same allele identically-by-decent [47, 48], an equivalent formulation supposes that the genetic effects can be modelled via **K**, the n × n kinship matrix, tabulating the measure of genetic relationship between the *i* and *j* participants in its *i,j* entry. Consider the vector form of the model for the *l* outcome with correlated errors of trait *l* = 1,2:

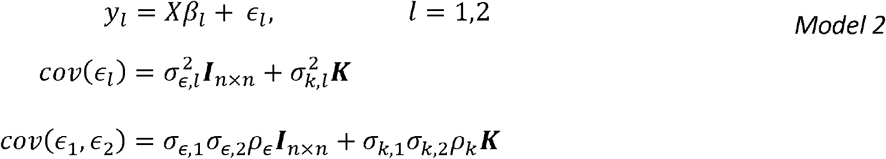

Now the genetic correlation can be estimated using mixed model with two traits. However, this is computationally demanding, especially for very large data sets. Recently, [49] discussed the Haseman-Elston regression for variance components estimation, and demonstrated that the genetic variance components estimator corresponding to the kinship matrix, if it is independent of all other correlation matrices in a model with potentially multiple sources of correlation (which holds here, because we only have a single correlation matrix, the kinship matrix) are given by 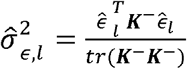, where *K*^−^is the kinship matrix with all diagonal values set to zero.

#### An estimator of genetic correlation between two phenotypes

We extend the Haseman-Elston approach for modelling the genetic correlations between two phenotypes. For the errors of persons *i* and *j*, and phenotypes 1 and 2, under *Model 2* we get:

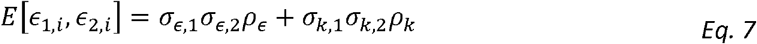

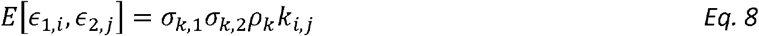

Suppose for now that 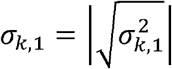 and 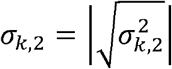 are known. For estimating the genetic correlation between the two phenotypes, we take all pairs 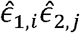 of residuals (estimating the error terms) after regression on mean-model covariates for *i* ≠ *j*, and regress them against the “covariate” *σ*_*k*,1_*σ*_*k*,2_*k*_*i,j*_. From properties of linear regression, we get:

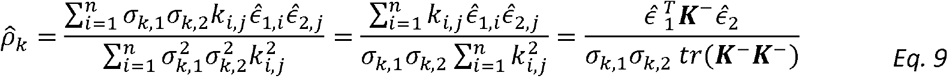

Now we can plug-in the estimators of *σ*_*k*,1_, *σ*_*k*,2_ to get:

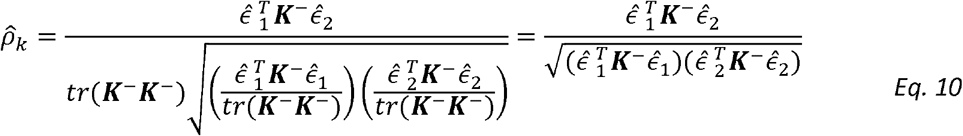

This estimator resembles that of the Pearson correlation parameter between the variables 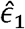 and 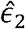, as can be seen if one replaces the matrix *K*^−^ by the identify matrix ***I***. Interestingly, this estimator does not involve the unknown variance parameters. It does include the estimated kinship parameters, which are treated as fixed.

#### Extension to multiple correlation matrices and generalization

We can use multiple relatedness matrices *A*,…,*K* with elements (*a*_*i,j*_..*k*_*i,j*_) indicating the measure of relatedness between the *i-th* and *j-th* participants in its *i,j* entry to model the variance. We can then estimate the variance components obtained from expressions of the form

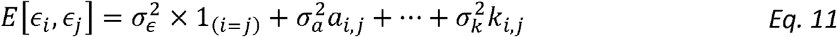

(corresponding to *Model 2* above) via a residual regression, i.e. by taking the vector all pairs of residuals 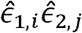 for all *i,j*. The HE design matrix, now re-defined (compared to [49]) to include rows corresponding to 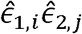 with *i* = *j*, is given by:

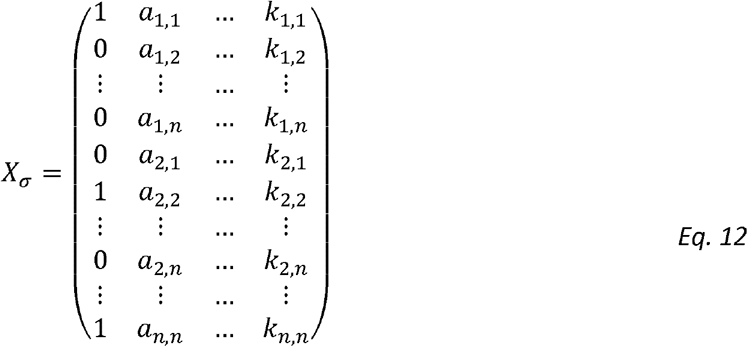

Similarly, the design matrix for estimating genetic correlation, obtained from expression of the form

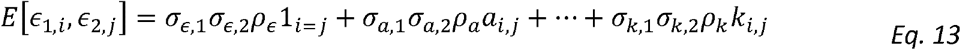

Can be written (if the variance parameters were known) as:

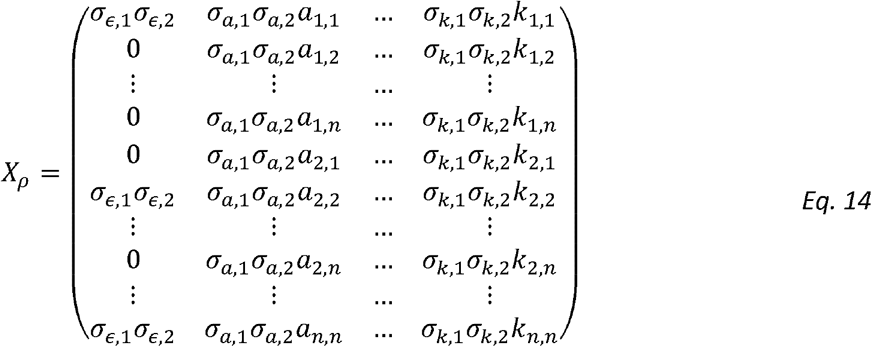

Noting that:

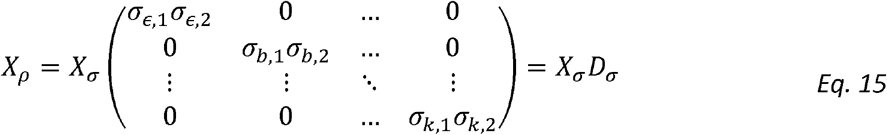

We get that:

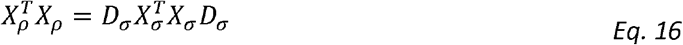

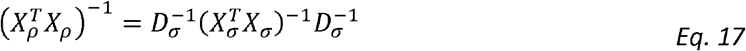

We also note the outcome matrices for estimating variance components and correlation parameters are:

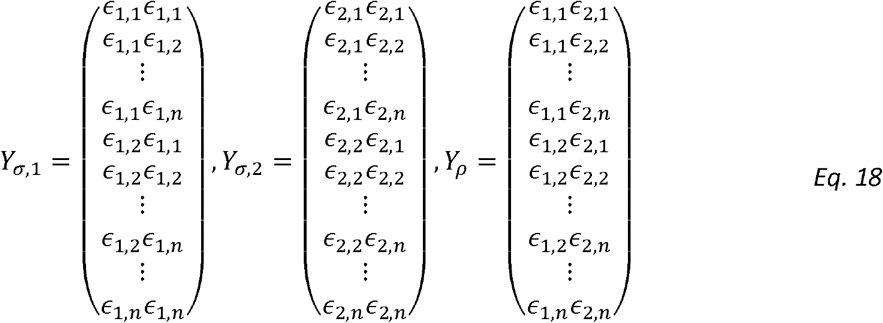

Therefore:

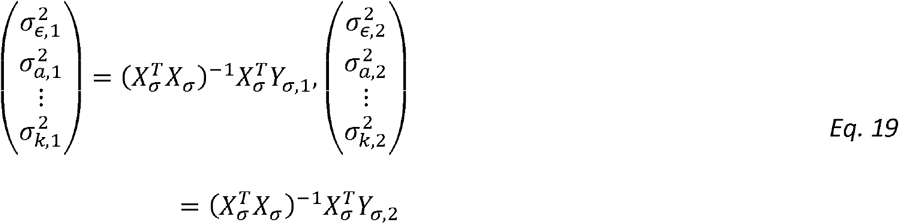

and:

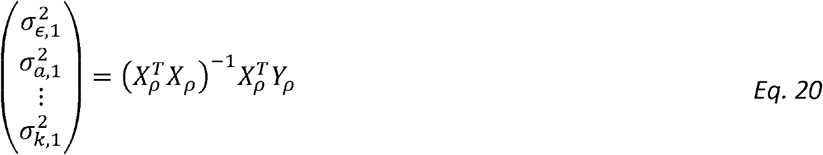

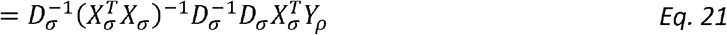

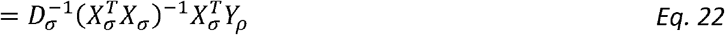

Because the *l*_th_ entry of *D*_*σ*_ is *σ*_1,*l*_ *σ*_2,*l*_ we have then for *ρ*_*l*_:

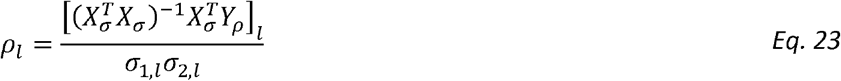

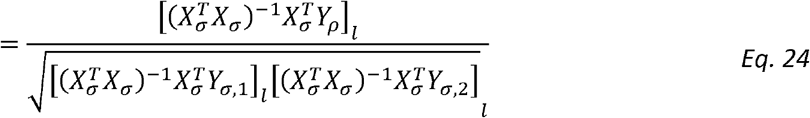

To prove that this is a generalized Pearson correlation, we only need to show that 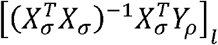 is a bilinear form, with the matrix completely defined by the *l*_th_ row of 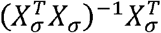. This is simple to see, because the entries of 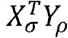 are:

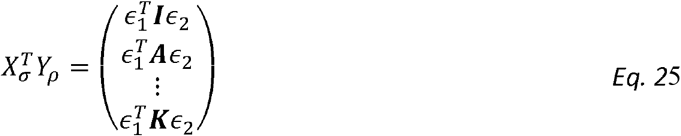

Similarly:

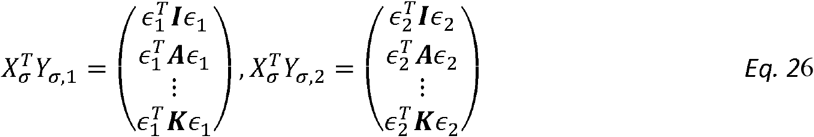

and the *l*_th_ row of 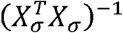 determines the weights in the following expression:

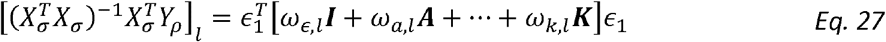

Thus for

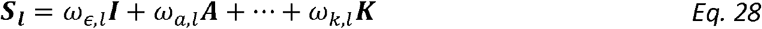

We get

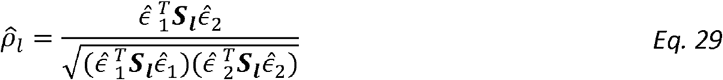

#### Deriving confidence intervals for estimated correlation coefficients

We propose two methods to compute confidence intervals for the estimated correlation coefficients. First, using the Fisher’s transformation, which was developed to estimate confidence intervals for the standard Pearson correlation coefficient, and second, using block bootstrap.

##### Confidence intervals using the Fisher’s transform

Fisher’s transformation converts the distribution of the correlation coefficients to a normal one and thus allows to calculate confidence intervals (and corresponding p-values) for the correlation coefficient using the values of the correlation coefficient and the sample size [50, 51]. Since we show that calculating genetic correlation is equal to calculating standard correlation for adjusted phenotypes (Eq. 29), the Fisher method is equally applicable for genetic correlation coefficient with the modification of plugging-in “effective sample size” to account for the modelled correlation structure between the two traits. Specifically, Fisher’s z-transformation of a correlation coefficient *ρ* is defined as:

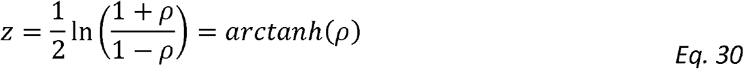

If the two variables for which the correlation is measured have a bivariate normal distribution and are independent and identically distributed, then *z* is approximately normally distributed with mean μ and standard error *σ* given by:

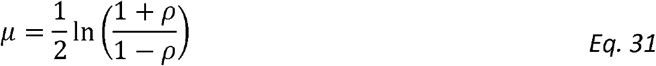

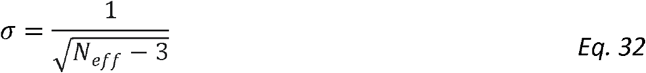

and *N*_*eff*_ being the effective sample size of our sample equal to:

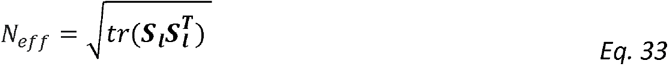

where ***S***_***l***_ is the weighted matrix described in *Eq. 28*. The coverage of this approach was verified in simulations via comparisons to the block Bootstrap method described below (see Supplementary Fig 1).

##### Confidence intervals using block Bootstrap

Multiple participants in our datasets are genetically related (and thus correlated), which violates the assumption of the standard bootstrap method. We thus performed the block bootstrap procedure to derive the confidence intervals and p-values as described in [52]. Briefly, related individuals were grouped into blocks (via 3^rd^ degree kinship) and the sampling procedure was at the level of blocks. Standard deviations and confidence intervals were calculated from the bootstrapped values that were Fisher’s transformed.

### 2.2. Heritability-Normalized Genetic Correlation

For simplicity, focus on the single correlation matrix settings (the derivation here naturally extends to multiple sources of correlation). Recall *Model 2 -* the phenotypic correlation coefficient ***R*** is equal to:

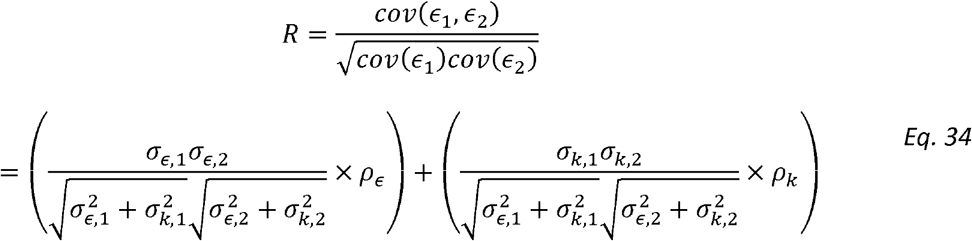

We then define Normalized Genetic Correlation (*ρ*_*Nk*_)as

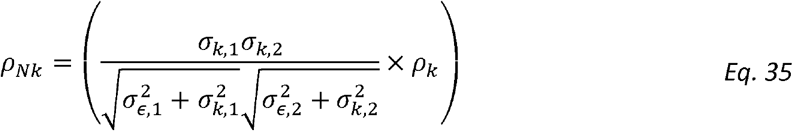

and Normalized Residual Correlation as 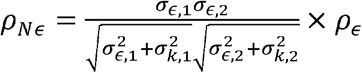.

This is a natural decomposition of phenotypic correlation into two components:

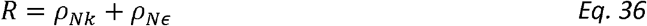

Unlike standard genetic correlation, *ρ*_*Nk*_ is the genetic correlation adjusted (normalized) for both traits’ heritabilities and variances and crucially it represents a fraction of the phenotypic correlation, normalized for the total phenotypic correlation, that is due to genetics. Thus, the normalized correlation terms are never larger than the phenotypic correlation *R*, because they sum to *R*.

### 2.3. Simulation studies

We studied the accuracy of the proposed method for estimating genetic correlations and for calculating confidence intervals in simulations. We used correlation matrices from the HCHS/SOL representing kinship and shared household to generate realistic correlation structures. In all simulations, data were generated by first sampling two uncorrelated error vectors (*ϵ*_1_, *ϵ*_2_)from a standard normal distribution. We next simulated the covariance structure according to our model:

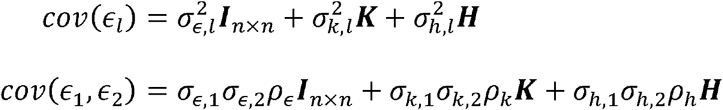

The matrix **K** represents kinship, and **H** represents shared household. All simulations were performed 1,000 times with different sample sizes (1000, 4000, and 7706, the latter is sample size of HCHS/SOL individuals in TOPMed freeze 8 which is the smallest subpopulation in this study) and values of *ρ*_*k*_ and *ρ*_*h*_ ranging from 0 to 1 in increments of 0.1. The variance components reported here were set to typical values for phenotypes from our dataset and equal to *σ* = (*σ*_*k*,1_ *σ*_*k*,2_, *σ*_*h*,1_, *σ*_*h*,2_)= (0.6,0.7,0.4,0.3) corresponding to e.g., HDL, Height, Fasting Glucose levels and Eosinophil counts. The confidence intervals and p-values were calculated via the Fisher transformation and block bootstrap methods as described above.

### 2.4. The Trans-Omics for Precision Medicine (TOPMed) Program

We used harmonized phenotype data from eight cohort studies participating in TOPMed (9) Freeze 8 [https://www.nhlbiwgs.org/topmed-whole-genome-sequencing-methods-freeze-8], which included 33,959 genotyped individuals from the Amish Study (n□= □1,105), JHS (n□= □2,807), FHS (n□= □3,658), HCHS/SOL (n=7,693), ARIC (n=7,479), CHS (n=3,482), MESA (n=4,665), and CARDIA (n=3,070) with available race/ethnic identification. Descriptions of each of these studies are provided in the Supplementary Materials. This dataset included 8,054 Black participants, 17,143 White participants and 8,762 participants of Hispanic/Latino descent. All participants provided informed consent and the study was approved by IRBs in each of the participating institutions. For TOPMed WGS data acquisition and QC report see ncbi.nlm.nih.gov/projects/gap/cgi□bin/GetPdf.cgi?id=phd006969.1. The phenotype harmonization was performed by TOPMed Data Coordinating Center (DCC) as described in [29]. The phenotype names and description, exclusion criteria and transformations are described in Supplementary Table 1. Phenotypes’ characteristics across race/ethnicities are reported in Supplementary Table 2. All analyses were adjusted for age, sex, study, and reported race/ethnicity as well as 5 first principal components (PCs) to adjust for population structure. The PCs, Kinship matrices, and unrelated individual pools were computed by TOPMed DCC via a robust pipeline [https://github.com/UW-GAC/analysis_pipeline] via a combination of KING [47], PC-AiR [53], and PC-Relate[48].

### 2.5. The Hispanic Community Health Study/Study of Latinos

The HCHS/SOL is a community-based cohort study of Hispanic individuals from four field centers across the US [30, 39] with almost 13,000 genotyped participants. A two-stage sampling scheme for participant selection was employed, with sampled community block units followed by households. Correlation matrices to model environmental variance due to households and community block units were generated so that the *i, j* entry of a given matrix was set to 1 if the *i* and *j* individuals live in the same household (or community block unit), and 0 otherwise. This study was approved by the institutional review boards at each field center, where all participants gave written informed consent. Genotyping and quality control for HCHS/SOL have been described in detail elsewhere[53]. In brief, DNA extracted from blood was genotyped on the HCHS Custom 15041502 array (Illumina Omni2.5M + custom content). Genotyping and downstream quality-control procedures yielded 2 232 944 genetic variants for genotyped HCHS/SOL participants. These were used for genotype imputation with the 1000 Genomes Project phase 3 reference panel [54] as previously described [55]. Variants with at least two copies of the minor allele and present in any of the four 1000 Genomes continental panels were imputed yielding about 50 million imputed variants prior to quality filtering. A subset of 7,693 individuals from HCHS/SOL participated in TOPMed and was used in the first part of this study (with the whole dataset used in the latter parts), with additional phenotypes that were not harmonized across other TOPMed studies. HCHS/SOL phenotype names and description are described in Supplementary Table 3. The number of participants with non-missing information per phenotype as well as means and standard deviations per phenotype per gender is reported in Supplementary Table 4. We computed genetic correlations between phenotypes from the following domains: anthropometric, blood pressure, lipids, blood cell counts, and inflammation markers. All analyses were adjusted for age, sex, sampling weights and five first principal components to account for population structure.

### 2.6. Heritability and Genetic/Environment Correlation estimation

The relatedness between individuals is modelled via a kinship (K) matrix, and an additional household matrix (H) for modelling environmental effects (available only for the HCHS/SOL cohort). Each phenotype was regressed on age, sex, sampling weights and 5 first principal components (and race/ethnicity and study for TOPMed combined cohorts) and the residuals were rank-normalized. We estimated the correlation coefficients corresponding to the relatedness matrices for all trait pairs by plugging in the normalized residuals to *Equation 29*. This was implemented via R scripts provided in GitHub repository [https://github.com/tamartsi/HE_Genetic_Correlation]. The genetic and environment variance components as well as the corresponding heritabilities were calculated via the GCTA software [31]. Following sensitivity analysis for the presence of related individuals in the cohorts (Supplementary Figure 2) we removed all individuals related at 3^rd^ degree or more for the calculation of heritabilities, however as we didn’t see any significant effects of relatives on the estimated genetic correlations (Supplementary Figure 2B), the relatives were kept in for genetic and environmental correlation coefficients estimation. Bootstrap was used to estimate standard deviations and confidence intervals. Visualizations were performed via the R packages igraph[56], qgraph[57], ggplot2[58] and corrplot[59] followed by Adobe Illustrator. The figures are based on uncorrected p-values. FDR-corrected p-values are provided in the Supplementary Data files.

### 2.7. Domain-Level enrichment analysis

We have calculated the enrichment of inter-domain correlation via a permutation approach. Specifically for 1000 repeats, we generated random connections between nodes in our correlation graph such that each node will receive a same number of connections as in the real dataset as well as keeping the overall number of connections identical. We then calculated the distribution of number of connections between each pair of domains and used it to obtain a domain enrichment p-value as follows:

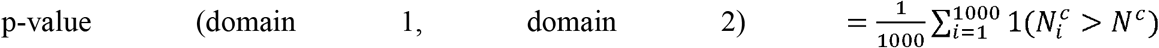

where *N*^*c*^ is the number of connections between domains 1 and 2, and 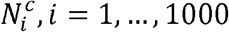 is the number of connections between domains 1 and 2 in the *i*th permutation. We considered two domains to be enriched if their enrichment p-value was < 0.05.

## Data Availability

TOPMed freeze 8 WGS data are available by application to dbGaP according to the study specific accessions specified in Supplementary Materials.

## 1. List of Abbreviations

BBJ: BioBank Japan
UKB: UK Biobank
GREML: genetic restricted maximum likelihood analysis
GWAS: genome-wide association study
LD: linkage disequilibrium
LDSC: linkage disequilibrium score regression
HE: Haseman-Elston regression
FDR: False Discovery Rate
TOPMed: Trans-Omics in Precision Medicine
HCHS/SOL: Hispanic Community Health Study / Study of Latinos
BMI: body-mass index
HDL: high-density lipoprotein
WBC: white blood cell counts
CRP: C-Reactive Protein
RBC: red blood cells
AHI: Apnea Hypopnea Index

## 5. Author Contributions

ME, MOG, and TS devised and developed the HE models and framework. ME and TS performed all the analyses, and summarized results in tables and figures. ME and TS conceptualized and drafted the manuscript. TS supervised the work for this manuscript. CI, HC, PSV, HX, AWM, XG, NF, BMP, SSR, JIR, DMLJ, MF, AC, NLHC, RSV, RH, RCS and SR designed data collection and/or TOPMed sample selection in the cohort they represent, formulated best-practices for data analysis for the same cohorts and participated in the manuscript preparation.

## 7. Data and Code availability

TOPMed freeze 8 WGS data are available by application to dbGaP according to the study specific accessions specified in Supplementary Materials. The code is available in GitHub repository [https://github.com/tamartsi/HE_Genetic_Correlation].

