## Supplementary Figures&Tables for "Genetic and environmental correlations between complex phenotypes differ by race/ethnicity and sex"

Elgart et al.

### Descriptions, acknowledgements, and ethics statements of studies participating in the analysis

#### HCHS/SOL

The Hispanic Community Health Study/Study of Latinos (dbGaP accession phs000810) is a community-based longitudinal cohort study of 16,415 self-identified Hispanic/Latino persons aged 18–74 years and selected from households in predefined census-block groups across four US field centers (in Chicago, Miami, the Bronx, and San Diego). The census-block groups were chosen to provide diversity among cohort participants with regard to socioeconomic status and national origin or background [1, 2]. The HCHS/SOL cohort includes participants who self-identified as having a Hispanic/Latino background; the largest groups are Central American (n = 1,730), Cuban (n = 2,348), Dominican (n = 1,460), Mexican (n = 6,471), Puerto Rican (n = 2,728), and South American (n = 1,068). The HCHS/SOL baseline clinical examination occurred between 2008 and 2011 and included comprehensive biological, behavioral, and sociodemographic assessments. Visit 2 took place between 2014 and 2017, which re-examined 11,623 participants from the baseline sample. Visit 3 has started in 2020 and will last 3 years. In addition to clinic visit, participants are contacted annually to assess clinical outcomes. The study was approved by the Institutional Review Boards at each participating institution and written informed consent was obtained from all participants.

*Ethics statement*: This study was approved by the institutional review boards (IRBs) at each field center, where all participants gave written informed consent, and by the Non-Biomedical IRB at the University of North Carolina at Chapel Hill, to the HCHS/SOL Data Coordinating Center. All IRBs approving the study are: Non-Biomedical IRB at the University of North Carolina at Chapel Hill. Chapel Hill, NC; Einstein IRB at the Albert Einstein College of Medicine of Yeshiva University. Bronx, NY; IRB at Office for the Protection of Research Subjects (OPRS), University of Illinois at Chicago. Chicago, IL; Human Subject Research Office, University of Miami. Miami, FL; Institutional Review Board of San Diego State University. San Diego, CA.

Acknowledgements: The Hispanic Community Health Study/Study of Latinos is a collaborative study supported by contracts from the National Heart, Lung, and Blood Institute (NHLBI) to the University of North Carolina (HHSN268201300001I / N01-HC-65233), University of Miami (HHSN268201300004I / N01-HC-65234), Albert Einstein College of Medicine (HHSN268201300002I / N01-HC-65235), University of Illinois at Chicago – HHSN268201300003I / N01-HC-65236 Northwestern Univ), and San Diego State University (HHSN268201300005I / N01-HC-65237). The following Institutes/Centers/Offices have contributed to the HCHS/SOL through a transfer of funds to the NHLBI: National Institute on Minority Health and Health Disparities, National Institute on Deafness and Other Communication Disorders, National Institute of Dental and Craniofacial Research, National Institute of Diabetes and Digestive and Kidney Diseases, National Institute of Neurological Disorders and Stroke, NIH Institution-Office of Dietary Supplements.

#### FHS

The Framingham Heart Study (dbGaP accession phs000007) began in 1948 with the recruitment of an original cohort of 5,209 men and women (mean age 44 years; 55 percent women). In 1971 a second generation of study participants was enrolled; this cohort (mean age 37 years; 52% women) consisted of 5,124 children and spouses of children of the original cohort. A third-generation cohort of 4,095 children of offspring cohort participants (mean age 40 years; 53 percent women) was enrolled in 2002-2005 and are seen every 4 to 8 years. Details of study designs for the three cohorts are summarized elsewhere [3–5]. At each clinic visit, a medical history was obtained, and participants underwent a physical examination. Only study participants consented for genetic data are included. FHS has been approved by the Boston University IRB

*Ethics statement*: The Framingham Heart Study was approved by the Institutional Review Board of the Boston University Medical Center. All study participants provided written informed consent.

Acknowledgements: The Framingham Heart Study (FHS) acknowledges the support of contracts NO1-HC-25195, HHSN268201500001I and 75N92019D00031 from the National Heart, Lung and Blood Institute and grant supplement R01 HL092577-06S1 for this research. We also acknowledge the dedication of the FHS study participants without whom this research would not be possible. Dr. Vasan is supported in part by the Evans Medical Foundation and the Jay and Louis Coffman Endowment from the Department of Medicine, Boston University School of Medicine.

#### ARIC

The Atherosclerosis Risk in Communities study (dbGaP accession phs000090) is a population-based prospective cohort study of cardiovascular disease sponsored by the NHLBI. ARIC included 15,792 individuals, predominantly European American and African American, aged 45-64 years at baseline (1987-89), chosen by probability sampling from four US communities. Cohort members completed three additional triennial follow-up examinations, a fifth exam in 2011-2013, a sixth exam in 2016-2017, and a seventh exam in 2018-2019. The ARIC study has been described in detail previously [6].

*Ethics statement*: All subjects provided informed consent and the study was approved by the Institutional Review Board (IRB) of the Johns Hopkins University School of Medicine.

Acknowledgements: The Atherosclerosis Risk in Communities study has been funded in whole or in part with Federal funds from the National Heart, Lung, and Blood Institute, National Institutes of Health, Department of Health and Human Services (contract numbers HHSN268201700001I, HHSN268201700002I, HHSN268201700003I, HHSN268201700004I and HHSN268201700005I). The authors thank the staff and participants of the ARIC study for their important contributions.

#### CHS

The Cardiovascular Health Study (dbGaP accession phs000287) is a population-based cohort study initiated by the NHLBI in 1987 to determine the risk factors for development and progression of cardiovascular disease (CVD) in older adults, with an emphasis on subclinical measures. The study recruited 5,888 adults aged 65 or older at entry in four U.S. communities and conducted extensive annual clinical exams between 1989-1999 along with semi-annual phone calls, events adjudication, and subsequent data analyses and publications. Additional data are collected by studies ancillary to CHS. In June 1990, four Field Centers (Sacramento, CA; Hagerstown, MD; Winston-Salem, NC; Pittsburgh, PA) completed the recruitment of 5201 participants. Between November 1992 and June 1993, an additional 687 African Americans were recruited using similar methods. Blood samples were drawn from all participants at their baseline examination and during follow-up clinic visits and DNA was subsequently extracted from available samples.

*Ethics statement*: All CHS participants provided informed consent, and the study was approved by the Institutional Review Board [or ethics review committee] of University Washington.

Acknowledgements: Cardiovascular Health Study: This research was supported by contracts HHSN268201200036C, HHSN268200800007C, HHSN268201800001C, N01HC55222, N01HC85079, N01HC85080, N01HC85081, N01HC85082, N01HC85083, N01HC85086, 75N92021D00006, and grants U01HL080295 and U01HL130114 from the National Heart, Lung, and Blood Institute (NHLBI), with additional contribution from the National Institute of Neurological Disorders and Stroke (NINDS). Additional support was provided by R01AG023629 from the National Institute on Aging (NIA). A full list of principal CHS investigators and institutions can be found at CHS-NHLBI.org. The content is solely the responsibility of the authors and does not necessarily represent the official views of the National Institutes of Health.

#### MESA

The Multi-Ethnic Study of Atherosclerosis (dbGaP accession phs000209) is a study of the characteristics of subclinical cardiovascular disease (disease detected non-invasively before it has produced clinical signs and symptoms) and the risk factors that predict progression to clinically overt cardiovascular disease or progression of the subclinical disease [7]. MESA consisted of a diverse, population-based sample of an initial 6,814 asymptomatic men and women aged 45-84. 38 percent of the recruited participants were white, 28 percent African American, 22 percent Hispanic, and 12 percent Asian, predominantly of Chinese descent. Participants were recruited from six field centers across the United States: Wake Forest University, Columbia University, Johns Hopkins University, University of Minnesota, Northwestern University and University of California - Los Angeles. Participants are being followed for identification and characterization of cardiovascular disease events, including acute myocardial infarction and other forms of coronary heart disease (CHD), stroke, and congestive heart failure; for cardiovascular disease interventions; and for mortality. The first examination took place over two years, from July 2000 - July 2002. It was followed by five examination periods that were 17-20 months in length. Participants have been contacted every 9 to 12 months throughout the study to assess clinical morbidity and mortality.

*Ethics statement*: All MESA participants provided written informed consent, and the study was approved by the Institutional Review Boards at The Lundquist Institute (formerly Los Angeles BioMedical Research Institute) at Harbor-UCLA Medical Center, University of Washington, Wake Forest School of Medicine, Northwestern University, University of Minnesota, Columbia University, and Johns Hopkins University.

Acknowledgements: MESA and the MESA SHARe project are conducted and supported by the National Heart, Lung, and Blood Institute (NHLBI) in collaboration with MESA investigators. Support for MESA is provided by contracts HHSN268201500003I, N01-HC-95159, N01-HC-95160, N01-HC-95161, N01-HC-95162, N01-HC-95163, N01-HC-95164, N01-HC-95165, N01-HC-95166, N01-HC-95167, N01-HC-95168, N01-HC-95169, UL1-TR-000040, UL1-TR-001079, UL1-TR-001420. MESA Family is conducted and supported by the National Heart, Lung, and Blood Institute (NHLBI) in collaboration with MESA investigators. Support is provided by grants and contracts R01HL071051, R01HL071205, R01HL071250, R01HL071251, R01HL071258, R01HL071259, and by the National Center for Research Resources, Grant UL1RR033176. The provision of genotyping data was supported in part by the National Center for Advancing Translational Sciences, CTSI grant UL1TR001881, and the National Institute of Diabetes and Digestive and Kidney Disease Diabetes Research Center (DRC) grant DK063491 to the Southern California Diabetes Endocrinology Research Center.

#### CARDIA

The Coronary Artery Risk Development in Young Adults study (dbGaP accession phs000285) is a prospective multicenter study with 5,115 adults Caucasian and African American participants of the age group 18-30 years at baseline, recruited from four centers at the baseline examination in 1985-1986 [8]. The recruitment was done from the total community in Birmingham, AL, from selected census tracts in Chicago, IL and Minneapolis, MN; and from the Kaiser Permanente health plan membership in Oakland, CA. Nine examinations have been completed in the years 0, 2, 5, 7, 10, 15, 20, 25 and 30, with high retention rates (91%, 86%, 81%, 79%, 74%, 72%, 72%, and 71%, respectively) and written informed consent was obtained in each visit.

*Ethics statement*: All CARDIA participants provided informed consent, and the study was approved by the Institutional Review Boards of the University of Alabama at Birmingham and the University of Texas Health Science Center at Houston.

Acknowledgements: The Coronary Artery Risk Development in Young Adults Study (CARDIA) is conducted and supported by the National Heart, Lung, and Blood Institute (NHLBI) in collaboration with the University of Alabama at Birmingham (HHSN268201800005I & HHSN268201800007I), Northwestern University (HHSN268201800003I), University of Minnesota (HHSN268201800006I), and Kaiser Foundation Research Institute (HHSN268201800004I). CARDIA was also partially supported by the Intramural Research Program of the National Institute on Aging (NIA) and an intra-agency agreement between NIA and NHLBI (AG0005).

#### JHS

The Jackson Heart Study (dbGaP accession phs000286) is a longitudinal investigation of genetic and environmental risk factors associated with the disproportionate burden of cardiovascular disease in African Americans [9, 10]. JHS is funded by the NHLBI and the National Institute on Minority Health and Health Disparities (NIMHD). JHS is an expansion of the ARIC study in its Jackson Field Center. At baseline, the JHS recruited 5306 African American residents of the Jackson Mississippi Metropolitan Statistical Area aged, approximately 6.6% of all African American adults aged 35-84 residing in the area. Participants were recruited via random sampling

(17% of participants), volunteers (30%), prior participants in the Atherosclerosis Risk in Communities (ARIC) study (31%), and secondary family members (22%). Among these participants, approximately 3400 gave consent that allows genetic research. JHS participants received three back-to-back clinical examinations (Exam 1, 2000-2004; Exam 2, 2005-2008; and Exam 3, 2009-2013), and a fourth clinical examination has started in 2020. Participants are also contacted annually by telephone to update personal and health information including vital status, interim medical events, hospitalizations, functional status and sociocultural information.

*Ethics statement*: The JHS study was approved by Jackson State University, Tougaloo College, and the University of Mississippi Medical Center IRBs, and all participants provided written informed consent.

Acknowledgements: The Jackson Heart Study (JHS) is supported and conducted in collaboration with Jackson State University (HHSN268201800013I), Tougaloo College (HHSN268201800014I), the Mississippi State Department of Health (HHSN268201800015I) and the University of Mississippi Medical Center (HHSN268201800010I, HHSN268201800011I and HHSN268201800012I) contracts from the National Heart, Lung, and Blood Institute (NHLBI) and the National Institute on Minority Health and Health Disparities (NIMHD). The authors also wish to thank the staffs and participants of the JHS.

Disclaimer: The views expressed in this manuscript are those of the authors and do not necessarily represent the views of the National Heart, Lung, and Blood Institute; the National Institutes of Health; or the U.S. Department of Health and Human Services.

#### CFS

The Cleveland Family Study (CFS) was designed to examine the genetic basis of sleep apnea in 2,534 African-American and European-American individuals from 356 families. Index probands with confirmed sleep apnea were recruited from sleep centers in northern Ohio, supplemented with additional family members and neighborhood control families [11]. Four visits occurred between 1990 and 2006; in the first 3, data were collected in participants' homes while the last occurred in a clinical research center (2000 - 2006). Measurements included sleep apnea monitoring, blood pressure, anthropometry, spirometry and other related phenotypes. Blood samples (overnight fasting, before bed and following an oral glucose tolerance test), nasal and oral ultrasound, and ECG were also obtained during the 4th exam. Institutional Review Board approval and signed informed consent was obtained for all participants.

*Ethics statement*: Cleveland Family Study was approved by the Institutional Review Board (IRB) of Case Western Reserve University and Mass General Brigham (formerly Partners HealthCare). Written informed consent was obtained from all participants.

Acknowledgements: The Cleveland Family Study has been supported in part by National Institutes of Health grants [R01-HL046380, KL2-RR024990, R35-HL135818, and R01-HL113338].

#### Amish

The Amish Complex Disease Research Program includes a set of large community-based studies focused largely on cardiometabolic health carried out in the Old Order Amish (OOA) community of Lancaster, Pennsylvania. Founders of the OOA population of Lancaster County, PA immigrated to the Colonies from Western Europe in the early 1700's. There are now over 30,000 OOA individuals in the Lancaster area, nearly all of whom can trace their ancestry back 12-14 generations to approximately 400 founders. Investigators at the University of Maryland School of Medicine have been studying the genetic determinants of cardiometabolic health in this population since 1993. To date, over 9,000 Amish adults have participated in one or more of our studies, of whom 1,100 are represented in the .TOPMed Consortium.

*Ethics statement*: All study protocols were approved by the institutional review board at the University of Maryland Baltimore. Informed consent was obtained from each study participant.

Acknowledgements: The TOPMed component of the Amish Research Program was supported by NIH grants R01 HL121007, U01 HL072515, and R01 AG18728. Email Rhea Cosentino for additional input.

#### TOPMed and CCDG acknowledgements

Molecular data for the Trans-Omics in Precision Medicine (TOPMed) program was supported by the National Heart, Lung and Blood Institute (NHLBI). Genome sequencing for “NHLBI TOPMed: Whole Genome Sequencing and Related Phenotypes in the Framingham Heart Study” (phs000974.v4.p3) was performed at the Broad Institute Genomics Platform (3R01HL092577-06S1, 3U54HG003067-12S2). Genome sequencing for “NHLBI TOPMed: The Jackson Heart Study” (phs000964.v1.p1) was performed at the Northwest Genomics Center (HHSN268201100037C). Genome sequencing for the “NHLBI TOPMed: The Atherosclerosis Risk in Communities Study” (phs001211.v3.p2) was performed at the Broad Institute Genomics Platform (3R01HL092577-06S1) and the Baylor College of Medicine Human Genome Sequencing Center (HHSN268201500015C, 3U54HG003273-12S2). Genome sequencing for “NHLBI TOPMed: Coronary Artery Risk Development in Young Adults Study” (phs001612.v1.p1) was performed at the Baylor College of Medicine Human Genome Sequencing Center (HHSN268201600033I). Genome sequencing for “NHLBI TOPMed: Cleveland Family Study” (phs000954.v3.p2) was performed at the Northwest Genomics Center (3R01HL098433-05S1, HHSN268201600032I). Genomics sequencing for “NHLBI TOPMed: Cardiovascular Health Study” (phs001368.v2.p1) was performed at the Baylor College of Medicine Human Genome Sequencing Center (3U54HG003273-12S2, HHSN268201500015C, HHSN268201600033I). Genome sequencing for “NHLBI TOPMed: Hispanic Community Health Study/Study of Latinos” (phs001395.v1.p1) was performed at the Baylor College of Medicine Human Genome Sequencing Center (HHSN268201600033I). Genome sequencing for “NHLBI TOPMed: Multi-Ethnic Study of Atherosclerosis” (phs001416.v2.p1) was performed at Broad Institute Genomics Platform (HHSN268201500014C, 3U54HG003067-13S1). WGS for “NHLBI TOPMed: Genetics of Cardiometabolic Health in the Amish” (phs000956) was performed at the Broad Institute of MIT and Harvard (3R01HL121007-01S1). Core support including centralized genomic read mapping and genotype calling, along with variant quality metrics and filtering were provided by the TOPMed

Informatics Research Center (3R01HL-117626-02S1; contract HHSN268201800002I). Core support including phenotype harmonization, data management, sample-identity QC, and general program coordination were provided by the TOPMed Data Coordinating Center (R01HL-120393; U01HL-120393; contract HHSN268201800001I). We gratefully acknowledge the studies and participants who provided biological samples and data for TOPMed. The Genome Sequencing Program (GSP) was funded by the National Human Genome Research Institute (NHGRI), the National Heart, Lung, and Blood Institute (NHLBI), and the National Eye Institute (NEI). The GSP Coordinating Center (U24 HG008956) contributed to cross-program scientific initiatives and provided logistical and general study coordination. The Centers for Common Disease Genomics (CCDG) program was supported by NHGRI and NHLBI, and whole genome sequencing was performed at the Baylor College of Medicine Human Genome Sequencing Center (UM1 HG008898 and R01HL059367).

### Supplementary Tables

#### Supplementary Table 1. Phenotypes harmonized across participating studies by the TOPMed DCC used in this study

| Code | | Description |
|---|---|---|
| annotated_sex_1 | | Biological sex |
| Race/ethnicity | *hispanic_or_latino_1* | Indicator of reported Hispanic or Latino ethnicity; only used samples where this agreed with “race_us_1” |
| | *race_us_1* | Reported race of participant according to the United States administrative definition of race; only used samples where this agreed with “hispanic_or_latino_1” |
| age_at_height_baseline_1 | | Age |
| height_baseline_1 | | Body height |
| bmi_baseline_1 | | Body mass index |
| antihypertensive_meds_1 | | Indicator for use of antihypertensive medication at the time of blood pressure measurement |
| bp_systolic_1 | | Resting systolic blood pressure from the upper arm in a clinical setting; only used samples if blood pressure lowering medications were not used (antihypertensive_meds_1) |
| bp_diastolic_1 | | Resting diastolic blood pressure from the upper arm in a clinical setting; only used samples if blood pressure lowering medications were not used (antihypertensive_meds_1) |

| | |
|---|---|
| *lipid_lowering_medication_1* | Indicates whether participant was taking any lipid-lowering medication at blood draw to measure lipids phenotypes |
| total_cholesterol_1 | Blood mass concentration of total cholesterol; only used samples where no lipid medication was used (lipid_lowering_medication_1) |
| triglycerides_1 | Blood mass concentration of triglycerides; only used samples where no lipid medication was used (lipid_lowering_medication_1) |
| hdl_1 | Blood mass concentration of high-density lipoprotein cholesterol; only used samples where no lipid medication was used (lipid_lowering_medication_1) |
| ldl_1 | Blood mass concentration of low-density lipoprotein cholesterol; only used samples where no lipid medication was used (lipid_lowering_medication_1) |
| hemoglobin_mcnc_bld_1 | Measurement of mass per volume, or mass concentration (mcnc), of hemoglobin in the blood (bld) |
| hematocrit_vfr_bld_1 | Measurement of hematocrit, the fraction of volume (vfr) of blood (bld) that is composed of red blood cells |
| rbc_ncnc_bld_1 | Count by volume, or number concentration (ncnc), of red blood cells in the blood (bld) |
| wbc_ncnc_bld_1 | Count by volume, or number concentration (ncnc), of white blood cells in the blood (bld) |
| basophil_ncnc_bld_1 | Count by volume, or number concentration (ncnc), of basophils in the blood (bld) |
| eosinophil_ncnc_bld_1 | Count by volume, or number concentration (ncnc), of eosinophils in the blood (bld) |
| neutrophil_ncnc_bld_1 | Count by volume, or number concentration (ncnc), of neutrophils in the blood (bld) |
| lymphocyte_ncnc_bld_1 | Count by volume, or number concentration (ncnc), of lymphocytes in the blood (bld) |
| monocyte_ncnc_bld_1 | Count by volume, or number concentration (ncnc), of monocytes in the blood (bld) |

| | |
|---|---|
| platelet_ncnc_bld_1 | Count by volume, or number concentration (ncnc), of platelets in the blood (bld) |
| mch_entmass_rbc_1 | Measurement of the average mass (entmass) of hemoglobin per red blood cell(rbc), known as mean corpuscular hemoglobin (MCH) |
| mchc_mcnc_rbc_1 | Measurement of the mass concentration (mcnc) of hemoglobin in a given volume of packed red blood cells (rbc), known as mean corpuscular hemoglobin concentration (MCHC) |
| mcv_entvol_rbc_1 | Measurement of the average volume (entvol) of red blood cells (rbc), known as mean corpuscular volume (MCV) |
| pmv_entvol_bld_1 | Measurement of the mean volume (entvol) of platelets in the blood (bld), known as mean platelet volume (MPV or PMV) |
| rdw_ratio_rbc_1 | Measurement of the ratio of variation in width to the mean width of the red blood cell (rbc) volume distribution curve taken at +/- 1 CV, known as red cell distribution width (RDW) |
| cd40_1 | Cluster of differentiation 40 ligand (CD40) concentration in blood. |
| crp_1 | C-reactive protein (CRP) concentration in blood |
| eselectin_1 | E-selectin concentration in blood. |
| icam1_1 | Intercellular adhesion molecule 1 (ICAM1) concentration in blood |
| il1_beta_1 | Interleukin 1 beta (IL1b) concentration in blood |
| il6_1 | Interleukin 6 (IL6) concentration in blood |
| il10_1 | Interleukin 10 (IL10) concentration in blood |
| il18_1 | Interleukin 18 (IL18) concentration in blood |
| isoprostane_8_epi_pgf2a_1 | Isoprostane 8-epi-prostaglandin F2 alpha (8-epi-PGF2a) concentration in urine |
| lppla2_act_1 | Activity of lipoprotein-associated phospholipase A2 (LP-PLA2), also known as platelet-activating factor acetylhydrolase, measured in blood |
| lppla2_mass_1 | Mass of lipoprotein-associated phospholipase A2 (LP-PLA2), also known as |

| | |
|---|---|
| | platelet-activating factor acetylhydrolase, measured in blood |
| mcp1_1 | Monocyte chemoattractant protein-1 (MCP1), also known as C-C motif chemokine ligand 2, concentration in blood |
| mmp9_1 | Matrix metalloproteinase 9 (MMP9) concentration in blood |
| mpo_1 | Myeloperoxidase (MPO) concentration in blood |
| opg_1 | Osteoprotegerin (OPG) concentration in blood |
| pselectin_1 | P-selectin concentration in blood. |
| tnfa_1 | Tumor necrosis factor alpha (TNFa) concentration in blood |
| tnfa_r1_1 | Tumor necrosis factor alpha receptor 1 (TNFa-R1) concentration in blood |
| tnfr2_1 | Tumor necrosis factor receptor 2 (TNFR2) concentration in blood |

#### Supplementary Table 2. Summary statistics of TOPMed phenotypes used in this study.

| | Black<br>(N=8054) | Hispanic/Latino<br>(N=8762) | White<br>(N=17143) | Overall<br>(N=33959) |
|---|---|---|---|---|
| **Age** | | | | |
| Mean (SD) | 51.5 (16.4) | 48.3 (14.3) | 51.4 (16.0) | 50.6 (15.8) |
| Median [Min, Max] | 53.0 [17.0, 93.0] | 49.0 [18.0, 86.0] | 52.0 [5.00, 98.0] | 52.0 [5.00, 98.0] |
| **Sex** | | | | |
| Male | 3231 (40.1%) | 3730 (42.6%) | 7833 (45.7%) | 14794 (43.6%) |
| Female | 4823 (59.9%) | 5032 (57.4%) | 9310 (54.3%) | 19165 (56.4%) |
| **bp_systolic_1** | | | | |
| Mean (SD) | 121 (18.5) | 121 (17.2) | 118 (17.1) | 119 (17.4) |
| Median [Min, Max] | 118 [73.0, 246] | 119 [77.0, 218] | 116 [67.0, 227] | 117 [67.0, 246] |
| Missing | 3433 (42.6%) | 1900 (21.7%) | 3323 (19.4%) | 8656 (25.5%) |
| **bp_diastolic_1** | | | | |
| Mean (SD) | 74.7 (11.4) | 73.3 (10.7) | 71.8 (10.2) | 72.7 (10.6) |
| Median [Min, Max] | 74.0 [0, 144] | 73.0 [40.0, 135] | 71.0 [18.0, 130] | 72.0 [0, 144] |
| Missing | 3433 (42.6%) | 1900 (21.7%) | 3323 (19.4%) | 8656 (25.5%) |
| **height_baseline_1** | | | | |
| Mean (SD) | 169 (9.47) | 163 (9.28) | 168 (9.64) | 167 (9.82) |
| Median [Min, Max] | 168 [125, 207] | 162 [132, 198] | 168 [123, 203] | 166 [123, 207] |
| **bmi_baseline_1** | | | | |

| | | | | |
|---|---|---|---|---|
| Mean (SD) | 29.8 (6.87) | 30.0 (6.27) | 26.3 (4.77) | 28.1 (6.01) |
| Median [Min, Max] | 28.8 [14.2, 91.8] | 29.1 [14.3, 70.3] | 25.6 [14.4, 58.1] | 27.1 [14.2, 91.8] |
| Missing | 5 (0.1%) | 10 (0.1%) | 11 (0.1%) | 26 (0.1%) |
| **total_cholesterol_1** | | | | |
| Mean (SD) | 201 (41.0) | 200 (43.4) | 207 (39.9) | 204 (41.3) |
| Median [Min, Max] | 198 [76.0, 470] | 197 [62.0, 526] | 204 [58.8, 594] | 201 [58.8, 594] |
| Missing | 3140 (39.0%) | 1554 (17.7%) | 3302 (19.3%) | 7996 (23.5%) |
| **triglycerides_1** | | | | |
| Mean (SD) | 107 (80.8) | 136 (121) | 124 (84.8) | 124 (96.2) |
| Median [Min, Max] | 90.0 [16.0, 2040] | 113 [20.0, 6370] | 104 [11.0, 1880] | 103 [11.0, 6370] |
| Missing | 3140 (39.0%) | 1554 (17.7%) | 3301 (19.3%) | 7995 (23.5%) |
| **hdl_1** | | | | |
| Mean (SD) | 52.5 (15.5) | 49.0 (13.1) | 52.8 (16.4) | 51.7 (15.5) |
| Median [Min, Max] | 50.0 [19.0, 162] | 47.0 [13.0, 141] | 50.0 [9.63, 156] | 49.0 [9.63, 162] |
| Missing | 3141 (39.0%) | 1555 (17.7%) | 3309 (19.3%) | 8005 (23.6%) |
| **ldl_1** | | | | |
| Mean (SD) | 128 (37.8) | 124 (36.4) | 130 (36.7) | 128 (36.9) |
| Median [Min, Max] | 125 [11.6, 361] | 122 [23.8, 417] | 128 [13.8, 505] | 126 [11.6, 505] |
| Missing | 3187 (39.6%) | 1689 (19.3%) | 3496 (20.4%) | 8372 (24.7%) |
| **hemoglobin_mcnc_bld_1** | | | | |
| Mean (SD) | 13.3 (1.60) | 13.8 (1.53) | 14.1 (1.34) | 13.8 (1.50) |
| Median [Min, Max] | 13.2 [3.00, 44.9] | 13.7 [4.30, 19.1] | 14.0 [6.70, 19.9] | 13.8 [3.00, 44.9] |
| Missing | 2490 (30.9%) | 346 (3.9%) | 7686 (44.8%) | 10522 (31.0%) |
| **hematocrit_vfr_bld_1** | | | | |
| Mean (SD) | 39.9 (4.38) | 42.0 (4.16) | 41.7 (3.94) | 41.4 (4.21) |
| Median [Min, Max] | 39.7 [11.9, 61.6] | 41.9 [18.7, 60.8] | 41.5 [19.7, 56.7] | 41.2 [11.9, 61.6] |
| Missing | 2491 (30.9%) | 346 (3.9%) | 7684 (44.8%) | 10521 (31.0%) |
| **rbc_ncnc_bld_1** | | | | |
| Mean (SD) | 4.61 (0.541) | 4.71 (0.457) | 4.56 (0.488) | 4.64 (0.493) |
| Median [Min, Max] | 4.57 [1.53, 7.05] | 4.69 [2.21, 7.00] | 4.53 [2.24, 6.81] | 4.62 [1.53, 7.05] |
| Missing | 3551 (44.1%) | 383 (4.4%) | 10620 (61.9%) | 14554 (42.9%) |
| **wbc_ncnc_bld_1** | | | | |
| Mean (SD) | 5.76 (1.99) | 6.55 (2.07) | 6.17 (1.85) | 6.21 (1.98) |
| Median [Min, Max] | 5.50 [1.60, 59.5] | 6.30 [1.80, 55.0] | 5.90 [1.50, 42.0] | 5.90 [1.50, 59.5] |
| Missing | 2724 (33.8%) | 772 (8.8%) | 7685 (44.8%) | 11181 (32.9%) |
| **basophil_ncnc_bld_1** | | | | |
| Mean (SD) | 0.0335 (0.0327) | 0.0333 (0.0376) | 0.0357 (0.0363) | 0.0341 (0.0361) |
| Median [Min, Max] | 0.0273 [0, 0.348] | 0.0240 [0, 0.345] | 0.0324 [0, 0.312] | 0.0284 [0, 0.348] |
| Missing | 4080 (50.7%) | 1031 (11.8%) | 11678 (68.1%) | 16789 (49.4%) |
| **eosinophil_ncnc_bld_1** | | | | |
| Mean (SD) | 0.149 (0.133) | 0.191 (0.185) | 0.180 (0.133) | 0.177 (0.158) |
| Median [Min, Max] | 0.112 [0, 1.94] | 0.145 [0, 2.72] | 0.151 [0, 1.68] | 0.139 [0, 2.72] |
| Missing | 3669 (45.6%) | 1018 (11.6%) | 11434 (66.7%) | 16121 (47.5%) |

| | | | | |
|---|---|---|---|---|
| **neutrophil_ncnc_bld_1** | | | | |
| Mean (SD) | 3.16 (1.44) | 3.67 (1.58) | 3.62 (1.33) | 3.53 (1.49) |
| Median [Min, Max] | 2.89 [0.100, 13.2] | 3.53 [0.0340, 26.7] | 3.40 [0.147, 20.8] | 3.32 [0.0340, 26.7] |
| Missing | 3509 (43.6%) | 1016 (11.6%) | 11315 (66.0%) | 15840 (46.6%) |
| **lymphocyte_ncnc_bld_1** | | | | |
| Mean (SD) | 2.02 (0.737) | 2.12 (0.778) | 1.80 (0.875) | 1.99 (0.812) |
| Median [Min, Max] | 1.92 [0.0510, 18.7] | 2.03 [0.270, 33.3] | 1.70 [0.0750, 37.6] | 1.90 [0.0510, 37.6] |
| Missing | 3508 (43.6%) | 1016 (11.6%) | 11313 (66.0%) | 15837 (46.6%) |
| **monocyte_ncnc_bld_1** | | | | |
| Mean (SD) | 0.372 (0.161) | 0.523 (0.191) | 0.437 (0.191) | 0.457 (0.194) |
| Median [Min, Max] | 0.356 [0, 1.40] | 0.498 [0, 2.86] | 0.415 [0, 2.14] | 0.437 [0, 2.86] |
| Missing | 3520 (43.7%) | 1016 (11.6%) | 11325 (66.1%) | 15861 (46.7%) |
| **platelet_ncnc_bld_1** | | | | |
| Mean (SD) | 255 (68.6) | 250 (66.6) | 245 (67.2) | 249 (67.4) |
| Median [Min, Max] | 250 [0, 940] | 245 [23.0, 874] | 238 [45.0, 1220] | 243 [0, 1220] |
| Missing | 2520 (31.3%) | 359 (4.1%) | 8186 (47.8%) | 11065 (32.6%) |
| **mch_entmass_rbc_1** | | | | |
| Mean (SD) | 29.0 (4.66) | 29.2 (2.24) | 30.9 (1.70) | 29.7 (2.98) |
| Median [Min, Max] | 29.2 [15.3, 293] | 29.5 [12.5, 41.1] | 30.9 [19.5, 41.4] | 29.9 [12.5, 293] |
| Missing | 3551 (44.1%) | 383 (4.4%) | 10621 (62.0%) | 14555 (42.9%) |
| **mchc_mcnc_rbc_1** | | | | |
| Mean (SD) | 33.3 (1.34) | 32.7 (1.43) | 33.8 (0.984) | 33.3 (1.33) |
| Median [Min, Max] | 33.3 [27.7, 100] | 32.9 [22.2, 45.2] | 33.8 [25.3, 51.5] | 33.4 [22.2, 100] |
| Missing | 2724 (33.8%) | 346 (3.9%) | 7686 (44.8%) | 10756 (31.7%) |
| **mcv_entvol_rbc_1** | | | | |
| Mean (SD) | 86.9 (7.12) | 89.3 (5.99) | 90.9 (4.52) | 89.3 (6.03) |
| Median [Min, Max] | 87.4 [53.2, 293] | 89.0 [54.0, 122] | 90.9 [57.9, 126] | 89.7 [53.2, 293] |
| Missing | 3524 (43.8%) | 383 (4.4%) | 10616 (61.9%) | 14523 (42.8%) |
| **pmv_entvol_bld_1** | | | | |
| Mean (SD) | 9.19 (0.934) | 8.86 (0.703) | 8.78 (0.985) | 9.00 (0.980) |
| Median [Min, Max] | 9.10 [6.30, 14.2] | 8.80 [8.00, 10.0] | 8.70 [6.30, 13.6] | 8.90 [6.30, 14.2] |
| Missing | 5543 (68.8%) | 8754 (99.9%) | 14924 (87.1%) | 29221 (86.0%) |
| **rdw_ratio_rbc_1** | | | | |
| Mean (SD) | 13.7 (1.38) | 13.8 (1.31) | 13.1 (0.969) | 13.6 (1.30) |
| Median [Min, Max] | 13.4 [11.1, 27.7] | 13.5 [11.4, 27.3] | 12.9 [11.1, 22.6] | 13.4 [11.1, 27.7] |
| Missing | 5543 (68.8%) | 1125 (12.8%) | 14923 (87.1%) | 21591 (63.6%) |
| **crp_1** | | | | |
| Mean (SD) | 4.89 (7.38) | 4.24 (7.25) | 3.44 (6.57) | 4.05 (7.02) |
| Median [Min, Max] | 2.53 [0.0410, 136] | 2.25 [0.120, 317] | 1.66 [0.00700, 217] | 2.04 [0.00700, 317] |
| Missing | 1918 (23.8%) | 228 (2.6%) | 6143 (35.8%) | 8289 (24.4%) |
| **icam1_1** | | | | |
| Mean (SD) | 199 (93.8) | 291 (92.2) | 248 (96.8) | 239 (98.6) |
| Median [Min, Max] | 172 [0.350, 900] | 284 [115, 900] | 244 [55.8, 1320] | 231 [0.350, 1320] |

| | | | | |
|---|---|---|---|---|
| Missing | 6637 (82.4%) | 8444 (96.4%) | 12385 (72.2%) | 27466 (80.9%) |
| **il6_1** | | | | |
| Mean (SD) | 1.98 (1.70) | 1.64 (1.18) | 1.84 (1.80) | 1.85 (1.73) |
| Median [Min, Max] | 1.52 [0.147, 23.4] | 1.30 [0.165, 10.7] | 1.39 [0.129, 44.2] | 1.40 [0.129, 44.2] |
| Missing | 6143 (76.3%) | 7933 (90.5%) | 11165 (65.1%) | 25241 (74.3%) |
| **lppla2_act_1** | | | | |
| Mean (SD) | 87.6 (57.9) | 145 (40.4) | 102 (65.7) | 103 (64.0) |
| Median [Min, Max] | 82.9 [9.48, 262] | 148 [20.4, 300] | 96.2 [12.0, 314] | 109 [9.48, 314] |
| Missing | 6625 (82.3%) | 8047 (91.8%) | 11616 (67.8%) | 26288 (77.4%) |
| **lppla2_mass_1** | | | | |
| Mean (SD) | 228 (101) | 186 (57.0) | 280 (114) | 262 (112) |
| Median [Min, Max] | 200 [53.2, 737] | 178 [77.4, 564] | 245 [67.1, 944] | 231 [53.2, 944] |
| Missing | 6646 (82.5%) | 8062 (92.0%) | 11628 (67.8%) | 26336 (77.6%) |

Mean, Median and percent of missing data for the phenotypes and covariates used in this study. All the traits are presented for the whole database as well as broken down by race/ethnicity (Black, White, and Hispanic/Latino).

#### Supplementary Table 3. Code and descriptions of HCHS/SOL phenotypes used in this study

| Code | Description |
|---|---|
| AHI | Apnea/Hypopnea Events (3% desat) |
| MinSpO2 | Minimum oxyhemoglobin saturation during sleep |
| AvgSpO2 | Mean oxygen saturation during sleep |
| SpO290 | Percent sleep time with oxygen saturation less than 90% |
| Height | Height |
| BMI | Body Mass Index (kg/m$^2$) |
| WaistHip | Waist to Hip Ratio |
| FEV1FVC | FEV1 to FVC Ratio (%) |
| FEV1 | Forced Expiratory Volume |
| FEVC | Forced Vital Capacity |
| PhysHlth | Aggregate Physical Health Scale |
| BrchIdx | Overall Ankle Brachial Index (occ. failure incl.) |
| FastInsl | Insulin, fasting (converted to mU/L) |
| OGTTInsl | Insulin, post OGTT (converted to mU/L) |
| eGFRnodemo | eGFR based on serum cystatin C w/o demographics |
| eGFRwdemo | eGFR based on serum cystatin C, serum creatinine, gender, age and race |
| HOMA | HOMA-IR index of Insulin Resistance |
| BCell | HOMA-BCELL index of Insulin Resistance |
| GlycHemo | Glycosylated Hemoglobin in SI units (mmol/mol) |
| ECGAbnorm_Mj | Major ECG Abnormalities |

| | |
|---|---|
| ECGAbnorm_Min | Minor ECG Abnormalities |
| EpSleep | Epworth Sleepiness Scale |
| SleepDur | Self-reported sleep duration (difference in bed and wake times) (hours) |
| Insom | Women's Health Initiative Insomnia Rating Scale |
| SysBP | Systolic Blood Pressure |
| DiasBP | Diastolic Blood Pressure |
| Arter | Mean arterial pressure |
| Pulse | Pulse pressure |
| WBC | White Blood Count (x10e9) |
| RBC | Red Blood Count (x10e12) |
| Hemoglob | Hemoglobin (g/dL) |
| Hemocrit | % Hematocrit |
| CorpVol | Mean Corpuscular Volume (fl) |
| MnCorpHemo | Mean Corpuscular Hemoglobin (pg) |
| MnCorpHemoConc | Mean Corpuscular Hemoglobin Concentration (g/dL) |
| RedCellDistWdth | % Red Cell Distribution Width |
| PlateletCnt | Platelet Count (x10e9) |
| NeutCnt | Neutrophil Count (x10e9) |
| LymphCnt | Lymphocyte Count (x10e9) |
| MonoCnt | Monocyte Count (x10e9) |
| EosCnt | Eosinophil Count (x10e9) |
| BasoCnt | Basophil Count (x10e9) |
| Chol | Total cholesterol (mg/dL) |
| Triglyc | Triglycerides (mg/dL) |
| HDLChol | HDL-cholesterol (mg/dL) |
| LDLChol | LDL-cholesterol (mg/dL) |
| FastGluc | Glucose, fasting (mg/dL) |
| OGTTGluc | Glucose, post OGTT (mg/dL) |
| GlycoHemo | % Glycosylated Hemoglobin |
| Creat | Creatinine (mg/dL) |
| UrineCreat | Urine creatinine, random (mg/dL) |
| UrineMicroAlb | Urine microalbumin, random (mg/dL) |
| AlbCreat | Albumin/creatinine ratio (mg/g) |
| Fe | Iron (ug/dL) |
| FeBindCap | Total Iron Binding Capacity (TIBC) (ug/dL) |
| TransSat | % Transferrin saturation |
| CReactProt | High-sensitivity C-Reactive Protein (mg/L) |
| HrtRt | Heart Rate |
| PRDur | PR duration |
| QRSDur | QRS duration |

| | |
|---|---|
| QTDur | QT duration |
| Sex | Sex |

#### Supplementary Table 4. Summary statistics of HCHS/SOL phenotypes used in this study.

| | Male (N=4476) | Female (N=3202) | Overall (N=7678) |
|---|---|---|---|
| **Age** | | | |
| Mean (SD) | 47.1 (13.7) | 46.0 (14.2) | 46.6 (13.9) |
| Median [Min, Max] | 48.0 [18.0, 76.0] | 48.0 [18.0, 75.0] | 48.0 [18.0, 76.0] |
| **AHI** | | | |
| Mean (SD) | 5.01 (9.82) | 9.20 (15.7) | 6.75 (12.7) |
| Median [Min, Max] | 1.43 [0, 112] | 3.33 [0, 142] | 2.09 [0, 142] |
| Missing | 562 (12.6%) | 423 (13.2%) | 985 (12.8%) |
| **MinSpO2** | | | |
| Mean (SD) | 87.5 (5.68) | 86.2 (6.52) | 86.9 (6.08) |
| Median [Min, Max] | 89.1 [43.3, 98.1] | 88.0 [44.9, 96.2] | 88.7 [43.3, 98.1] |
| Missing | 550 (12.3%) | 417 (13.0%) | 967 (12.6%) |
| **AvgSpO2** | | | |
| Mean (SD) | 96.5 (0.861) | 96.3 (1.12) | 96.4 (0.984) |
| Median [Min, Max] | 96.8 [84.0, 99.4] | 96.6 [77.3, 98.2] | 96.7 [77.3, 99.4] |
| Missing | 550 (12.3%) | 417 (13.0%) | 967 (12.6%) |
| **SpO290** | | | |
| Mean (SD) | 0.635 (2.51) | 1.28 (4.15) | 0.902 (3.31) |
| Median [Min, Max] | 0.0330 [0, 87.1] | 0.0788 [0, 54.1] | 0.0502 [0, 87.1] |
| Missing | 550 (12.3%) | 417 (13.0%) | 967 (12.6%) |
| **Height** | | | |
| Mean (SD) | 157 (6.30) | 170 (6.96) | 163 (9.31) |
| Median [Min, Max] | 157 [132, 189] | 170 [135, 198] | 162 [132, 198] |
| Missing | 10 (0.2%) | 3 (0.1%) | 13 (0.2%) |
| **BMI** | | | |
| Mean (SD) | 30.8 (6.85) | 29.2 (5.61) | 30.1 (6.41) |
| Median [Min, Max] | 29.7 [14.3, 70.3] | 28.6 [14.9, 55.9] | 29.1 [14.3, 70.3] |
| Missing | 16 (0.4%) | 8 (0.2%) | 24 (0.3%) |
| **WaistHip** | | | |
| Mean (SD) | 0.899 (0.0736) | 0.954 (0.0716) | 0.922 (0.0778) |
| Median [Min, Max] | 0.900 [0.520, 1.31] | 0.953 [0.550, 1.25] | 0.923 [0.520, 1.31] |
| Missing | 10 (0.2%) | 15 (0.5%) | 25 (0.3%) |
| **FEV1FVC** | | | |
| Mean (SD) | 80.6 (7.31) | 78.7 (8.23) | 79.8 (7.76) |
| Median [Min, Max] | 81.4 [17.5, 100] | 79.8 [28.7, 100] | 80.8 [17.5, 100] |

| | | | |
|---|---|---|---|
| Missing | 284 (6.3%) | 162 (5.1%) | 446 (5.8%) |
| **FEV1** | | | |
| Mean (SD) | 2400 (592) | 3340 (788) | 2800 (824) |
| Median [Min, Max] | 2400 [0, 4920] | 3360 [0, 5790] | 2720 [0, 5790] |
| Missing | 226 (5.0%) | 140 (4.4%) | 366 (4.8%) |
| **FEVC** | | | |
| Mean (SD) | 2970 (686) | 4240 (869) | 3500 (989) |
| Median [Min, Max] | 2970 [0, 15100] | 4240 [0, 7380] | 3390 [0, 15100] |
| Missing | 226 (5.0%) | 140 (4.4%) | 366 (4.8%) |
| **PhysHlth** | | | |
| Mean (SD) | 47.2 (10.7) | 49.5 (9.52) | 48.2 (10.3) |
| Median [Min, Max] | 49.9 [11.5, 76.0] | 52.5 [7.70, 74.7] | 51.3 [7.70, 76.0] |
| Missing | 49 (1.1%) | 35 (1.1%) | 84 (1.1%) |
| **BrchIdx** | | | |
| Mean (SD) | 1.04 (0.122) | 1.09 (0.148) | 1.06 (0.135) |
| Median [Min, Max] | 1.03 [0.525, 2.52] | 1.09 [0, 2.34] | 1.06 [0, 2.52] |
| Missing | 1660 (37.1%) | 1293 (40.4%) | 2953 (38.5%) |
| **FastInsl** | | | |
| Mean (SD) | 13.6 (11.9) | 13.4 (16.4) | 13.5 (14.0) |
| Median [Min, Max] | 10.8 [0.559, 296] | 10.4 [0.559, 726] | 10.6 [0.559, 726] |
| Missing | 12 (0.3%) | 18 (0.6%) | 30 (0.4%) |
| **OGTTInsl** | | | |
| Mean (SD) | 93.9 (80.5) | 78.1 (78.8) | 87.2 (80.2) |
| Median [Min, Max] | 70.9 [2.57, 815] | 54.0 [1.50, 660] | 64.0 [1.50, 815] |
| Missing | 892 (19.9%) | 585 (18.3%) | 1477 (19.2%) |
| **eGFRnodemo** | | | |
| Mean (SD) | 110 (27.7) | 101 (23.8) | 106 (26.5) |
| Median [Min, Max] | 108 [5.77, 277] | 102 [4.89, 235] | 105 [4.89, 277] |
| Missing | 35 (0.8%) | 35 (1.1%) | 70 (0.9%) |
| **eGFRwdemo** | | | |
| Mean (SD) | 101 (20.7) | 100 (20.3) | 101 (20.5) |
| Median [Min, Max] | 102 [3.37, 177] | 103 [3.21, 163] | 103 [3.21, 177] |
| Missing | 35 (0.8%) | 36 (1.1%) | 71 (0.9%) |
| **HOMA** | | | |
| Mean (SD) | 3.63 (4.48) | 3.71 (4.98) | 3.66 (4.70) |
| Median [Min, Max] | 2.58 [0.104, 179] | 2.62 [0.115, 201] | 2.59 [0.104, 201] |
| Missing | 12 (0.3%) | 19 (0.6%) | 31 (0.4%) |
| **BCell** | | | |
| Mean (SD) | 150 (179) | 129 (136) | 141 (163) |
| Median [Min, Max] | 122 [2.67, 8280] | 100 [2.29, 5330] | 114 [2.29, 8280] |
| Missing | 19 (0.4%) | 20 (0.6%) | 39 (0.5%) |
| **GlycHemo** | | | |
| Mean (SD) | 40.8 (13.1) | 41.3 (15.0) | 41.0 (13.9) |

| | | | |
|---|---|---|---|
| Median [Min, Max] | 38.0 [17.0, 185] | 38.0 [13.0, 148] | 38.0 [13.0, 185] |
| Missing | 18 (0.4%) | 14 (0.4%) | 32 (0.4%) |
| **ECGAbnorm_Mj** | | | |
| Mean (SD) | 0.0869 (0.282) | 0.114 (0.318) | 0.0982 (0.298) |
| Median [Min, Max] | 0 [0, 1.00] | 0 [0, 1.00] | 0 [0, 1.00] |
| Missing | 47 (1.1%) | 27 (0.8%) | 74 (1.0%) |
| **ECGAbnorm_Min** | | | |
| Mean (SD) | 0.370 (0.483) | 0.563 (0.496) | 0.450 (0.498) |
| Median [Min, Max] | 0 [0, 1.00] | 1.00 [0, 1.00] | 0 [0, 1.00] |
| Missing | 47 (1.1%) | 27 (0.8%) | 74 (1.0%) |
| **EpSleep** | | | |
| Mean (SD) | 5.63 (4.85) | 5.91 (4.91) | 5.75 (4.88) |
| Median [Min, Max] | 5.00 [0, 24.0] | 5.00 [0, 24.0] | 5.00 [0, 24.0] |
| Missing | 96 (2.1%) | 56 (1.7%) | 152 (2.0%) |
| **SleepDur** | | | |
| Mean (SD) | 7.94 (1.49) | 7.81 (1.45) | 7.88 (1.47) |
| Median [Min, Max] | 8.00 [3.00, 13.5] | 7.79 [3.00, 13.0] | 7.93 [3.00, 13.5] |
| Missing | 245 (5.5%) | 178 (5.6%) | 423 (5.5%) |
| **Insom** | | | |
| Mean (SD) | 8.19 (5.73) | 6.65 (5.35) | 7.55 (5.63) |
| Median [Min, Max] | 7.00 [0, 20.0] | 5.00 [0, 20.0] | 6.00 [0, 20.0] |
| Missing | 139 (3.1%) | 107 (3.3%) | 246 (3.2%) |
| **SysBP** | | | |
| Mean (SD) | 122 (19.5) | 126 (16.5) | 123 (18.5) |
| Median [Min, Max] | 118 [76.0, 231] | 124 [86.0, 229] | 121 [76.0, 231] |
| Missing | 3 (0.1%) | 1 (0.0%) | 4 (0.1%) |
| **DiasBP** | | | |
| Mean (SD) | 73.6 (10.9) | 75.4 (11.1) | 74.3 (11.0) |
| Median [Min, Max] | 73.0 [41.0, 122] | 75.0 [43.0, 123] | 74.0 [41.0, 123] |
| Missing | 4 (0.1%) | 4 (0.1%) | 8 (0.1%) |
| **Arter** | | | |
| Mean (SD) | 89.6 (12.7) | 92.3 (11.9) | 90.7 (12.5) |
| Median [Min, Max] | 88.3 [55.3, 151] | 91.3 [61.3, 158] | 89.7 [55.3, 158] |
| Missing | 4 (0.1%) | 4 (0.1%) | 8 (0.1%) |
| **Pulse** | | | |
| Mean (SD) | 47.9 (14.2) | 50.9 (11.8) | 49.2 (13.3) |
| Median [Min, Max] | 45.0 [4.00, 128] | 49.0 [25.0, 120] | 47.0 [4.00, 128] |
| Missing | 4 (0.1%) | 4 (0.1%) | 8 (0.1%) |
| **WBC** | | | |
| Mean (SD) | 6.66 (2.00) | 6.45 (1.98) | 6.57 (1.99) |
| Median [Min, Max] | 6.40 [1.80, 31.8] | 6.20 [2.00, 25.5] | 6.40 [1.80, 31.8] |
| Missing | 258 (5.8%) | 233 (7.3%) | 491 (6.4%) |
| **RBC** | | | |

| | | | |
|---|---|---|---|
| Mean (SD) | 4.54 (0.354) | 5.01 (0.424) | 4.73 (0.449) |
| Median [Min, Max] | 4.54 [2.89, 6.51] | 5.03 [2.21, 7.00] | 4.71 [2.21, 7.00] |
| Missing | 31 (0.7%) | 32 (1.0%) | 63 (0.8%) |
| **Hemoglob** | | | |
| Mean (SD) | 13.0 (1.23) | 14.9 (1.21) | 13.8 (1.53) |
| Median [Min, Max] | 13.1 [4.30, 18.8] | 15.0 [6.30, 19.1] | 13.7 [4.30, 19.1] |
| Missing | 31 (0.7%) | 32 (1.0%) | 63 (0.8%) |
| **Hemocrit** | | | |
| Mean (SD) | 40.2 (3.39) | 44.8 (3.53) | 42.1 (4.13) |
| Median [Min, Max] | 40.2 [19.4, 55.8] | 44.9 [19.0, 60.8] | 42.0 [19.0, 60.8] |
| Missing | 31 (0.7%) | 32 (1.0%) | 63 (0.8%) |
| **CorpVol** | | | |
| Mean (SD) | 88.8 (6.29) | 89.8 (5.54) | 89.2 (6.01) |
| Median [Min, Max] | 89.0 [54.0, 122] | 90.0 [60.0, 121] | 89.0 [54.0, 122] |
| Missing | 31 (0.7%) | 32 (1.0%) | 63 (0.8%) |
| **MnCorpHemo** | | | |
| Mean (SD) | 28.7 (2.35) | 29.8 (1.88) | 29.1 (2.24) |
| Median [Min, Max] | 29.1 [12.5, 40.1] | 29.9 [17.6, 41.1] | 29.4 [12.5, 41.1] |
| Missing | 31 (0.7%) | 32 (1.0%) | 63 (0.8%) |
| **MnCorpHemoConc** | | | |
| Mean (SD) | 32.3 (1.42) | 33.2 (1.32) | 32.7 (1.45) |
| Median [Min, Max] | 32.5 [22.2, 43.2] | 33.3 [23.6, 37.4] | 32.8 [22.2, 43.2] |
| Missing | 31 (0.7%) | 32 (1.0%) | 63 (0.8%) |
| **RedCellDistWdth** | | | |
| Mean (SD) | 13.9 (1.43) | 13.5 (1.07) | 13.8 (1.31) |
| Median [Min, Max] | 13.6 [11.4, 27.3] | 13.3 [11.5, 24.0] | 13.5 [11.4, 27.3] |
| Missing | 32 (0.7%) | 32 (1.0%) | 64 (0.8%) |
| **PlateletCnt** | | | |
| Mean (SD) | 269 (67.3) | 227 (57.4) | 252 (66.6) |
| Median [Min, Max] | 264 [26.0, 874] | 224 [36.0, 707] | 247 [26.0, 874] |
| Missing | 36 (0.8%) | 33 (1.0%) | 69 (0.9%) |
| **NeutCnt** | | | |
| Mean (SD) | 3.77 (1.60) | 3.56 (1.60) | 3.68 (1.60) |
| Median [Min, Max] | 3.60 [0.100, 26.7] | 3.40 [0, 22.6] | 3.50 [0, 26.7] |
| Missing | 258 (5.8%) | 231 (7.2%) | 489 (6.4%) |
| **LymphCnt** | | | |
| Mean (SD) | 2.19 (0.702) | 2.07 (0.681) | 2.14 (0.696) |
| Median [Min, Max] | 2.10 [0.500, 6.80] | 2.00 [0.300, 6.30] | 2.00 [0.300, 6.80] |
| Missing | 258 (5.8%) | 232 (7.2%) | 490 (6.4%) |
| **MonoCnt** | | | |
| Mean (SD) | 0.502 (0.172) | 0.577 (0.334) | 0.533 (0.255) |
| Median [Min, Max] | 0.500 [0, 2.90] | 0.500 [0, 14.8] | 0.500 [0, 14.8] |
| Missing | 258 (5.8%) | 231 (7.2%) | 489 (6.4%) |

| | | | |
|---|---|---|---|
| **EosCnt** | | | |
| Mean (SD) | 0.179 (0.163) | 0.294 (4.22) | 0.227 (2.72) |
| Median [Min, Max] | 0.100 [0, 2.70] | 0.200 [0, 230] | 0.100 [0, 230] |
| Missing | 260 (5.8%) | 231 (7.2%) | 491 (6.4%) |
| **BasoCnt** | | | |
| Mean (SD) | 0.0216 (0.0433) | 0.0267 (0.0469) | 0.0237 (0.0449) |
| Median [Min, Max] | 0 [0, 0.300] | 0 [0, 0.300] | 0 [0, 0.300] |
| Missing | 266 (5.9%) | 237 (7.4%) | 503 (6.6%) |
| **Chol** | | | |
| Mean (SD) | 200 (43.6) | 197 (45.6) | 199 (44.5) |
| Median [Min, Max] | 196 [82.0, 526] | 194 [62.0, 587] | 195 [62.0, 587] |
| Missing | 3 (0.1%) | 3 (0.1%) | 6 (0.1%) |
| **Triglyc** | | | |
| Mean (SD) | 125 (79.3) | 154 (158) | 137 (119) |
| Median [Min, Max] | 107 [22.0, 1310] | 123 [20.0, 6370] | 113 [20.0, 6370] |
| Missing | 3 (0.1%) | 3 (0.1%) | 6 (0.1%) |
| **HDLChol** | | | |
| Mean (SD) | 52.3 (13.1) | 45.0 (12.3) | 49.2 (13.3) |
| Median [Min, Max] | 51.0 [16.0, 135] | 43.0 [13.0, 141] | 47.0 [13.0, 141] |
| Missing | 3 (0.1%) | 4 (0.1%) | 7 (0.1%) |
| **LDLChol** | | | |
| Mean (SD) | 123 (37.2) | 122 (37.3) | 123 (37.3) |
| Median [Min, Max] | 120 [34.0, 334] | 120 [22.0, 417] | 120 [22.0, 417] |
| Missing | 48 (1.1%) | 98 (3.1%) | 146 (1.9%) |
| **FastGluc** | | | |
| Mean (SD) | 93.8 (9.78) | 97.2 (9.58) | 95.2 (9.84) |
| Median [Min, Max] | 92.0 [52.0, 125] | 96.0 [53.0, 125] | 94.0 [52.0, 125] |
| Missing | 424 (9.5%) | 352 (11.0%) | 776 (10.1%) |
| **OGTTGluc** | | | |
| Mean (SD) | 127 (42.6) | 119 (42.7) | 123 (42.8) |
| Median [Min, Max] | 119 [38.0, 389] | 110 [29.0, 329] | 115 [29.0, 389] |
| Missing | 891 (19.9%) | 571 (17.8%) | 1462 (19.0%) |
| **GlycoHemo** | | | |
| Mean (SD) | 5.88 (1.20) | 5.92 (1.37) | 5.90 (1.28) |
| Median [Min, Max] | 5.60 [3.70, 19.1] | 5.60 [3.30, 15.7] | 5.60 [3.30, 19.1] |
| Missing | 18 (0.4%) | 14 (0.4%) | 32 (0.4%) |
| **Creat** | | | |
| Mean (SD) | 0.749 (0.257) | 0.997 (0.447) | 0.852 (0.370) |
| Median [Min, Max] | 0.730 [0.270, 11.9] | 0.950 [0.440, 15.1] | 0.810 [0.270, 15.1] |
| Missing | 3 (0.1%) | 3 (0.1%) | 6 (0.1%) |
| **UrineCreat** | | | |
| Mean (SD) | 127 (75.0) | 165 (80.9) | 143 (79.7) |
| Median [Min, Max] | 116 [7.00, 550] | 155 [5.00, 598] | 133 [5.00, 598] |

| | | | |
|---|---|---|---|
| Missing | 19 (0.4%) | 16 (0.5%) | 35 (0.5%) |
| **UrineMicroAlb** | | | |
| Mean (SD) | 35.1 (201) | 58.4 (346) | 44.9 (272) |
| Median [Min, Max] | 9.00 [2.00, 5790] | 9.00 [2.00, 10900] | 9.00 [2.00, 10900] |
| Missing | 210 (4.7%) | 106 (3.3%) | 316 (4.1%) |
| **AlbCreat** | | | |
| Mean (SD) | 35.6 (290) | 49.7 (395) | 41.6 (338) |
| Median [Min, Max] | 7.37 [1.41, 12600] | 5.48 [1.02, 16800] | 6.57 [1.02, 16800] |
| Missing | 209 (4.7%) | 106 (3.3%) | 315 (4.1%) |
| **Fe** | | | |
| Mean (SD) | 80.1 (32.0) | 99.8 (34.5) | 88.3 (34.4) |
| Median [Min, Max] | 78.0 [9.00, 320] | 96.0 [15.0, 317] | 85.0 [9.00, 320] |
| Missing | 3 (0.1%) | 4 (0.1%) | 7 (0.1%) |
| **FeBindCap** | | | |
| Mean (SD) | 326 (50.0) | 308 (42.1) | 319 (47.7) |
| Median [Min, Max] | 321 [171, 595] | 305 [173, 614] | 314 [171, 614] |
| Missing | 3 (0.1%) | 4 (0.1%) | 7 (0.1%) |
| **TransSat** | | | |
| Mean (SD) | 25.3 (10.9) | 32.9 (11.9) | 28.5 (11.9) |
| Median [Min, Max] | 25.0 [2.00, 99.0] | 32.0 [3.00, 99.0] | 27.0 [2.00, 99.0] |
| Missing | 3 (0.1%) | 4 (0.1%) | 7 (0.1%) |
| **CReactProt** | | | |
| Mean (SD) | 4.97 (8.19) | 3.35 (6.23) | 4.29 (7.48) |
| Median [Min, Max] | 2.84 [0.120, 317] | 1.75 [0.120, 128] | 2.26 [0.120, 317] |
| Missing | 3 (0.1%) | 3 (0.1%) | 6 (0.1%) |
| **HrtRt** | | | |
| Mean (SD) | 64.5 (10.8) | 63.2 (15.6) | 64.0 (13.0) |
| Median [Min, Max] | 63.0 [39.0, 357] | 62.0 [38.0, 416] | 63.0 [38.0, 416] |
| Missing | 55 (1.2%) | 32 (1.0%) | 87 (1.1%) |
| **PRDur** | | | |
| Mean (SD) | 155 (20.8) | 160 (23.6) | 157 (22.1) |
| Median [Min, Max] | 153 [80.0, 400] | 156 [0, 421] | 154 [0, 421] |
| Missing | 54 (1.2%) | 47 (1.5%) | 101 (1.3%) |
| **QRSDur** | | | |
| Mean (SD) | 89.3 (10.7) | 97.9 (13.3) | 92.9 (12.6) |
| Median [Min, Max] | 88.0 [52.0, 171] | 96.0 [0, 196] | 92.0 [0, 196] |
| Missing | 47 (1.1%) | 27 (0.8%) | 74 (1.0%) |
| **QTDur** | | | |
| Mean (SD) | 419 (28.0) | 410 (29.7) | 415 (29.1) |
| Median [Min, Max] | 418 [246, 550] | 408 [310, 563] | 414 [246, 563] |
| Missing | 47 (1.1%) | 28 (0.9%) | 75 (1.0%) |

Mean, Median and percent of missing data for the phenotypes and covariates used in this study. All the traits are presented for the whole database as well as broken down by gender.

### Supplementary Figures

#### Supplementary Figure 1. Comparison of simulations between our method, the ground truth and the GREML approach.

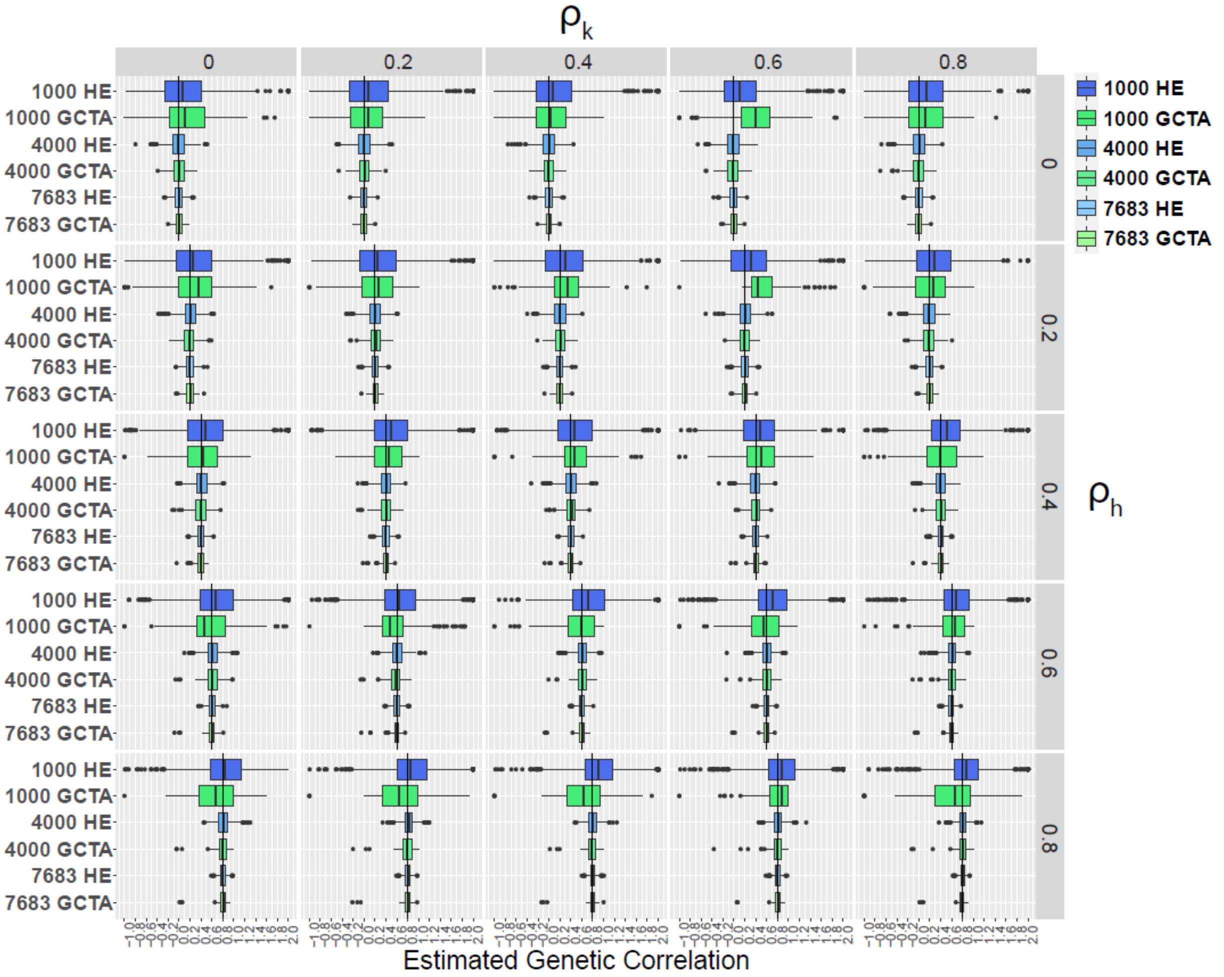

Two relatedness matrices were used to simulate phenotypes with known correlation coefficients ($\rho_h, \rho_k$). Each phenotype was simulated 1000 times in 1000, 4000 and 7683 people. Shown here are the boxplots of distributions of estimated $\rho_k$ for both our closed-form HE approach (blue colors) as well as the gold-standard REML method GCTA (green colors) [12].

#### Supplementary Figure 2. Sensitivity analysis of the presence of relatives in cohort on heritability and genetic correlation

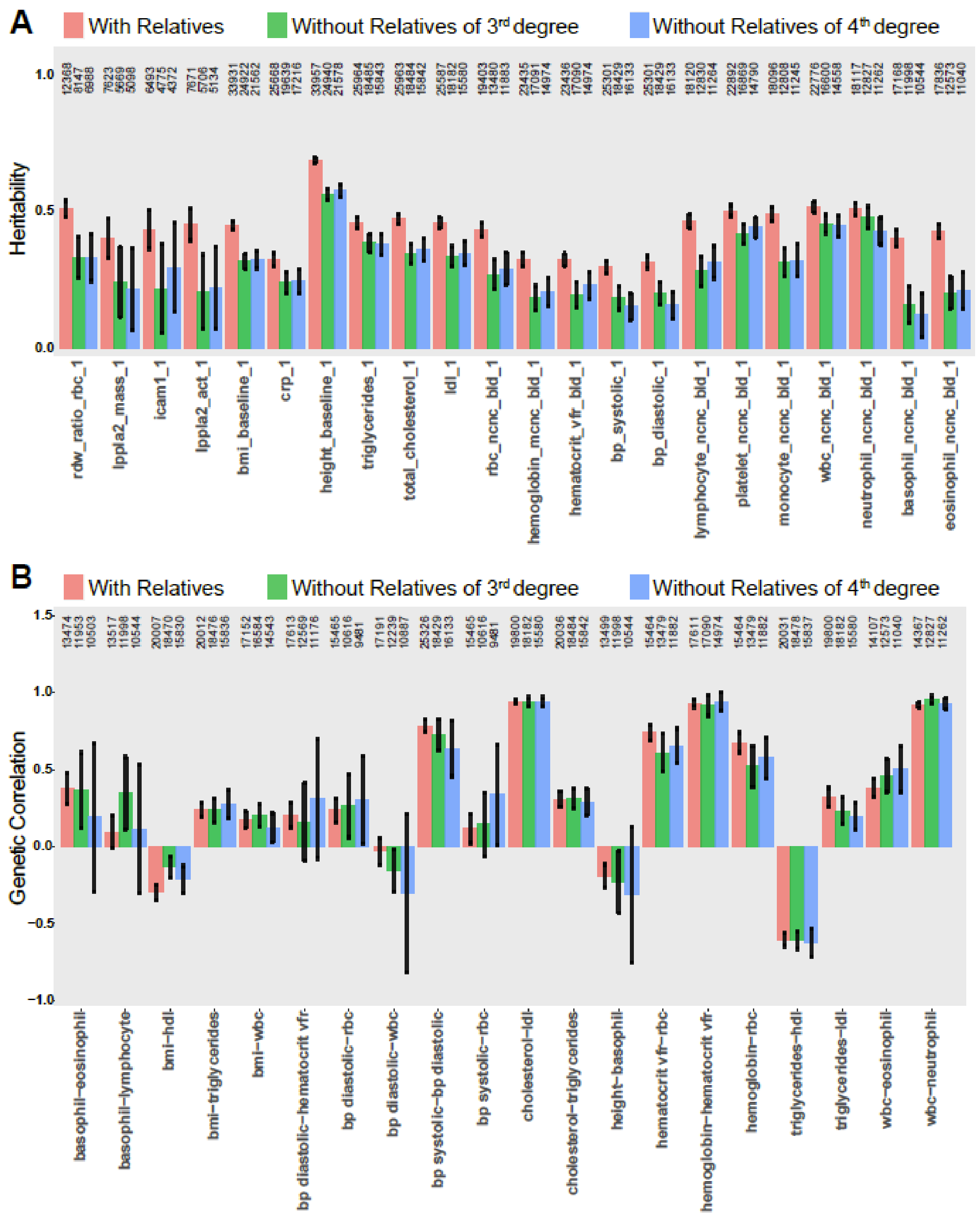

**(A, B)** Analysis of the TOPMed cohort with the relatives present (pink), with relatives of 3rd degree and more distant removed (green), and with relatives of 4th degree and more distant removed (blue) for selected examples of phenotypes with regard to **(A)** Heritability and **(B)** Genetic correlations ($\rho_k$).

#### Supplementary Figure 3. Certain genetic correlations are race/ethnicity-specific.

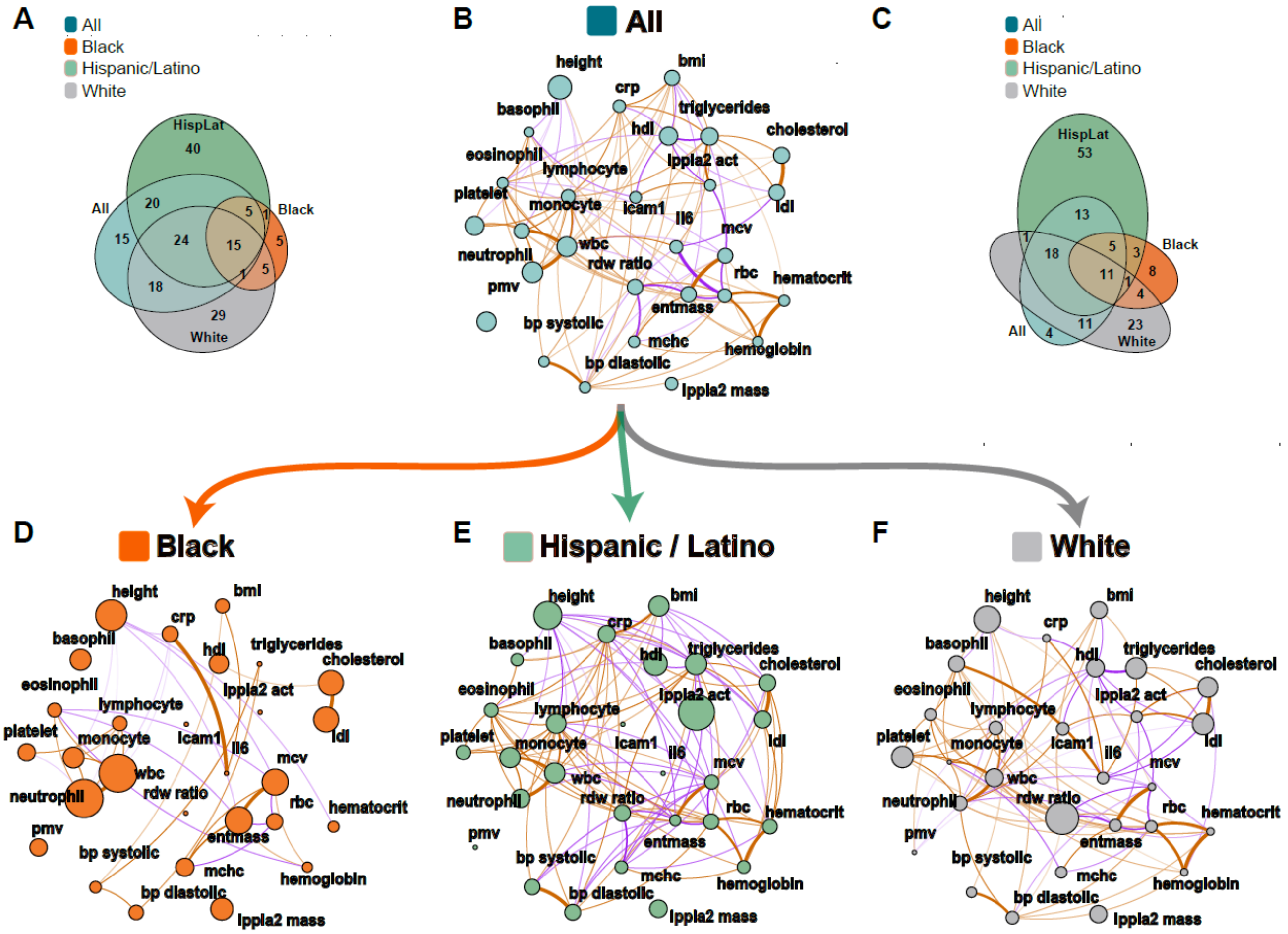

**(B, D-F)** Correlation plots where each phenotype is represented by a node and the correlations are represented by connections (edges) between nodes. The size of the node is proportional to the phenotype heritability. The thickness of the edge is proportional to the strength of correlation and the color represents magnitude: orange represents positive and purple negative correlation. **(B)** Genetic correlations ($\rho_k$) between the 28 phenotypes in the combined TOPMed dataset (p-value < 0.05; $|\rho_k| > 0.05$) **(D, E, F)** Genetic correlations ($\rho_k$) between the 28 phenotypes in the race/ethnic-specific subsets of the TOPMed dataset - Black (**D**, orange), Hispanic/Latino (**E**, marine) and White (**F**, grey). **(A, C)** Venn diagrams depicting the overlap in significant phenotype-pairs between Black (orange), Hispanic/Latino (marine) and White (grey) and a combined dataset (navy blue) for genetic correlation **(A)**, and normalized genetic correlation **(C)**.

#### Supplementary Figure 4. Genetic and environmental correlations and heritabilities differ by sex in Hispanics/Latinos.

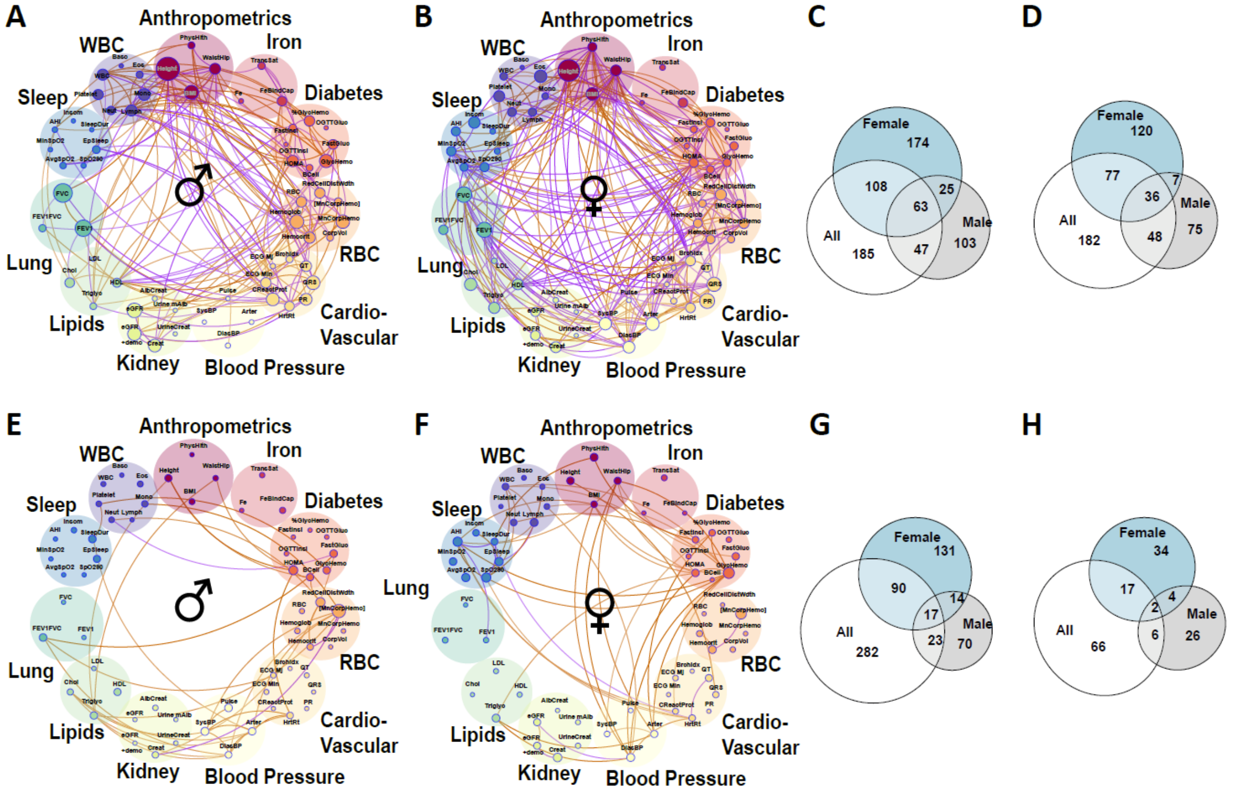

**(A-D)** Correlation plots where each phenotype is represented by a node and the correlations are represented by connections (edges) between nodes. The size of the node is proportional to the phenotype heritability. The thickness of the edge is proportional to the strength of correlation and the color represents magnitude: orange represents positive and purple negative correlation. Shown are normalized genetic correlations ($\rho_{Nk}$) between the 61 phenotypes in the extended HCHS/SOL dataset (p-value < 0.05; $|\rho_{Nk}| > 0.05$). Correlations and heritabilities as measured in males **(A, E)** and females **(B, F).** The top panels represent genetic interactions **(A, B)** and the bottom panels **(E, F)** represent the environmental interactions. (C, D, G, H) Venn diagrams depicting the overlap in significant phenotype-pairs between Males, Females and a combined dataset (All) for genetic correlation **(C)**, household correlation **(G)**, normalized genetic correlation **(D)** and normalized household correlation **(H)**.

### Supplementary Figure 5. Genetic correlations in the White group stratified by sex.

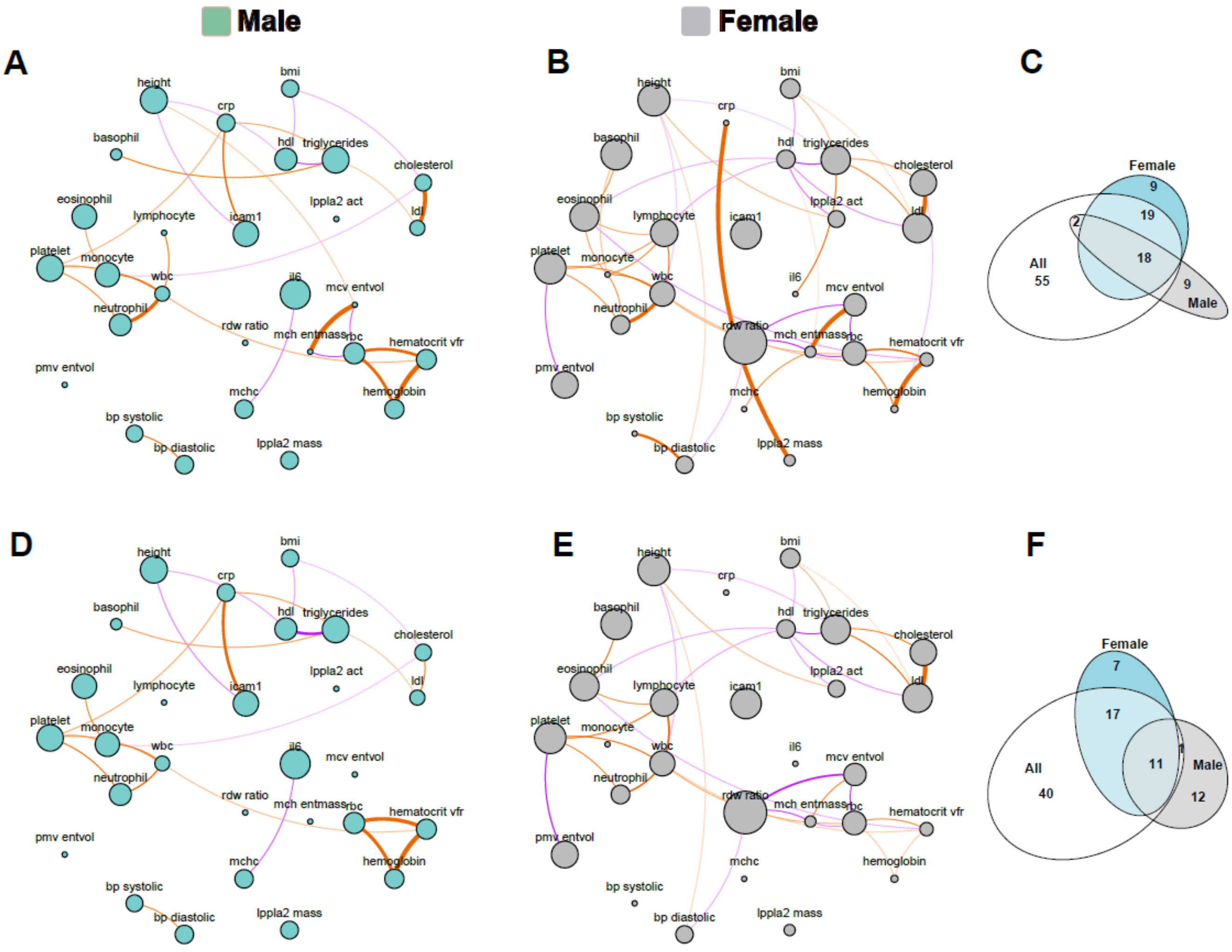

**(A-D)** Correlation plots where each phenotype is represented by a node and the correlations are represented by connections (edges) between nodes. The size of the node is proportional to the phenotype heritability. The thickness of the edge is proportional to the strength of correlation and the color represents magnitude: orange represents positive and purple negative correlation. Shown are genetic correlations ($\rho_k$) **(A, D)** between the 18 phenotypes in the TOPMed dataset (p-value < 0.05; $|\rho_k| > 0.05$); and normalized genetic correlations ($\rho_{Nk}$) **(A, D)** between the 18 phenotypes in the TOPMed dataset (p-value < 0.05; $|\rho_{Nk}| > 0.05$). The dataset was stratified by males **(A, D)** and females **(B, E).** Venn diagrams depicting the overlap in significant phenotype-pairs **(C,F)** between males, females and a combined dataset (All) for genetic correlation **(C)**, and normalized household correlation **(F)**.
